# Anti-Spike IgG4 and Fc Effector Responses: The Impact of SARS-CoV-2 Vaccine Platform–Specific Priming and Immune Imprinting

**DOI:** 10.1101/2025.03.19.25324267

**Authors:** Raj Kalkeri, Mingzhu Zhu, Shane Cloney-Clark, Anand Parekh, Drew Gorinson, Zhaohui Cai, Miranda R. Cai, Soham Mahato, Gordon Chau, Tara M. Babu, Anna Wald, Pradhipa Ramanathan, L. Carissa Aurelia, Kevin John Selva, Anthony M. Marchese, Louis Fries, Lisa M. Dunkle, Amy W. Chung, Joyce S. Plested

## Abstract

Proportional increases in anti-Spike (S) IgG4 associated with decreased Fc effector functions have been reported following repeated mRNA, but not recombinant protein-based (rS) (NVX-CoV2373, Novavax), SARS-CoV-2 vaccination. We demonstrate the first evidence of a negative correlation between anti-S IgG4 and neutralizing antibody (nAb) as well as antibody-dependent Fc effector functions. Priming with two NVX-CoV2373 vaccines followed by a third dose was associated with higher IgG1 and IgG3, lower IgG4, higher nAb titers and Fc effector functions versus mRNA. Immune imprinting of anti-S IgG4 and nAbs, and Fc effector function imprinting after mRNA priming was observed. This effect was partially overcome by updated XBB.1.5 protein subunit vaccination, but not ancestral vaccine strains. We establish correlation of anti-S IgG4 responses to reduced nAbs and Fc effector functions and demonstrate the impact of additional booster vaccination on subsequent immune response and Fc effector functions in the context of ancestral and XBB.1.5 strains.

## Main

Immunoglobulin G (IgG) subclass 4 (IgG4) typically accounts for a small proportion of total IgG (~0–5%), but the proportion of IgG4 within a given specific immune response is known to increase after repeated exposure to certain antigens and may exhibit characteristics related to immune tolerance^1–3^. Repeated mRNA SARS-CoV-2 vaccination has been associated with proportional elevations of SARS-CoV-2 Spike (S)-specific IgG4^4–19^. By contrast, increases in anti-S IgG4 have not been observed following repeated vaccination with adenoviral vectored (ChAdOx1 nCoV-19, Oxford/AstraZeneca) or Matrix-M™ adjuvanted recombinant SARS-CoV-2 S (rS) protein (NVX-CoV2373, Novavax, Inc.) vaccines ^5,7,10,11,14,19^. These studies together suggest that anti-S IgG4 class switch is induced by COVID-19 vaccines based on mRNA platforms. The precise mechanisms underlying this difference remain unclear and more robust data are required^20^. Nonetheless, IgG4 is known to exhibit an approximately 10-fold lower affinity for Fcγ receptors, such as FcγRIIa and FcγRIIIa, compared to other IgG subclasses, and to be a less efficient activator of complement^21–24^. Thus, increased IgG4 has the potential to interfere with effector immune response, such as Fc-receptor mediated antibody-dependent cellular phagocytosis (ADCP), cellular cytotoxicity (ADCC), complement deposition (ADCD), and natural killer (NK) cell activation^3,24–26^, implying a potentially adverse effect on some immune functions. The impact of IgG4 is nuanced and dependent on levels and specificities of other IgGs; whether IgG class switch to vaccine-induced anti-S IgG4 affects immunity to SARS-CoV-2 remains unknown as is the long-term clinical relevance of this phenomenon.

Several types of repeatedly administered vaccines targeting a variety of different diseases, including pertussis, human immunodeficiency virus (HIV) infection, and malaria, have been shown to induce vaccine antigen-specific IgG4 responses^20^. These observations are nuanced, complicating the understanding of mechanisms that may drive the vaccine IgG4 response. For example, antigen-specific IgG4 induction has been observed by the acellular but not whole-cell pertussis vaccines, and alum-adjuvanted DPALFA HIV vaccine induced significantly higher IgG4 responses compared to MLPA + QS-21 (saponin) DIP-10 PALFQ HIV vaccine^20^. It is hypothesized that a combination of high–frequency dosing intervals, excessive and persistent antigen exposure, and/or unique cytokine activation patterns drive IgG4 class switch^1–3,20^. In the context of COVID-19 vaccination, Irrgang et al. demonstrated that administration of a third dose of the mRNA vaccine (BNT162b2, Pfizer Inc. or mRNA-1273, Moderna, Inc.) elicited increases in anti-S IgG4 as a percentage of total anti-S IgG^8^. Furthermore, the shift in the IgG profile appears to persist over time and higher IgG4 is shown to be correlated with impaired Fc-mediated effector functioning in vitro^8^. In agreement with previous studies, we recently reported that repeated mRNA COVID-19 vaccination was associated with an increase in S-specific IgG4; by contrast, IgG4 class switch was not observed following priming and subsequent doses (four total vaccine doses) of the protein-based rS COVID-19 vaccine^27^. SARS-CoV-2–specific IgG3, an IgG subclass known to induce potent neutralization^28^, was notably higher after the NVX-CoV2373 rS vaccine (>10x vs. mRNA recipients)^27^. In contrast to regimens featuring mRNA vaccines only, vaccine regimens of NVX-CoV2373 only, or NVX-CoV2373 as a booster to prior mRNA immunizations, demonstrated higher levels of Fcγ effector activities^27^. Here, we show an inverse correlation between anti-S IgG4 levels with measurable elements of functional anti-SARS-CoV-2 immunity and further characterize the S-specific IgG subclasses induced by rS and mRNA COVID-19 vaccines. The work also provides evidence of the unique influence of COVID-19 vaccine type–specific priming on subsequent immune responses and investigates the related potential impact of the rS vaccine based on newer viral COVID-19 variants.

## Results

### Study Populations

The clinical samples included in this analysis came from participants in several clinical trials who had received different COVID-19 vaccine types and regimens. Study designs and demographic characteristics varied somewhat among the four study populations and are specified in **Table 1**. Participants from 2019nCoV-307 (N=19) had previously received three doses (3x) of ancestral sequence mRNA (Pfizer or Moderna) and received a 4^th^ dose with the ancestral protein-based rS vaccine (NVX-CoV2373). Participants from 2019nCoV-301 (N=18) had previously received three doses (3x) of ancestral-sequence protein-based rS followed by a 4^th^ dose of the same protein-based vaccine. Participants from the University of Washington VRC443 study (N=30) had previously received two doses (2x) of ancestral sequence protein-based rS followed by a 3^rd^ dose with a commercial Pfizer mRNA vaccine. Finally, participants from 2019nCoV-313 (N=29) had previously received three or more doses of the ancestral-sequence commercial mRNA vaccine followed by a dose of the updated XBB.1.5 protein-based rS vaccine (NVX-CoV2601). All study groups were 70–90% White, with a small predominance of women, and median age in the fifth decade, the sole exception being the older age group of 2019nCoV-313 subjects **(Table 1)**.

**Table 1.** Study demography, including vaccination regime, vaccine type, sample size, age, sex, race, serostatus (anti-NP), and time interval.

| *Parameter | 2019nCoV-307 (Group A) | 2019nCoV-301 (Group B) | VRC443 (Group C) | 2019nCoV-313 (Group D) |
| --- | --- | --- | --- | --- |
| **Priming Vaccinations**<br>Two (2x) or<br>Three (3x) Doses | Ancestral mRNA<br>BNT162b2 (Pfizer), OR<br>mRNA-1273 (Moderna)<br>(3x) | Ancestral Protein-based<br>NVX-CoV2373 (Novavax)<br>(3x) | Ancestral Protein-based<br>NVX-CoV2373<br>(2x) | Ancestral mRNA<br>BNT162b2 (Pfizer), OR<br>mRNA-1273 (Moderna)<br>($\geq 3x$) |
| **Additional Dose Vaccine**<br>Single Dose (1x) | Ancestral Protein-based<br>NVX-CoV2373 (Novavax) | Ancestral Protein-based<br>NVX-CoV2373 (Novavax) | Ancestral mRNA<br>BNT162b2 (Pfizer) | XBB.1.5 Strain Protein-based<br>NVX-CoV2601 (Novavax) |
| **Number of Volunteers**<br>N | 19 | 18 | 30 | 29 |
| **Race**<br>N (%) | White, 17 (89.5);<br>African American, 1 (5.3);<br>Asian, 1 (5.3) | White, 12 (66.7);<br>African American, 2 (11.1);<br>American Indian, 2 (11.1);<br>Asian, 2 (11.1) | White, 21 (70.0);<br>Asian, 3 (10.0);<br>>1 Race, 3 (10.0);<br>Other, 3 (10.0) | White, 23 (79.3);<br>American Indian Or Alaska Native, 1 (3.4);<br>Asian, 1 (3.4);<br>Black Or African American, 1 (3.4);<br>Native Hawaiian Or Other Pacific Islander, 1 (3.4);<br>Not Reported, 1 (3.4);<br>Unknown, 1 (3.4) |
| **Age in Years**<br>Median (IQR range) | 41 (37 to 46) | 53.5 (43 to 68) | 47 (29 to 67) | 65 (58 to 72) |
| **Sex**<br>Female N (%) | 10 (52.6%) | 12 (66.7%) | 17 (57) | 18 (62.1) |
| **Serostatus** (anti-NP<br>seropositive, seronegative) | 8, 10 | 8, 10 | 5, 19 | 19, 10 |
| **Sex**<br>Male N (%) | 9 (47.4%) | 6 (33.3%) | 13 (43) | 11 (37.9) |
| **Time Interval Between 3<sup>rd</sup> to 4<sup>th</sup> Immunization**<br>Median Days (IQR range) | 238 (217 to 259) | 258 (253 to 266) | 291 (216 to 315) | 309 (296 to 352) |

\*Some differences in parameter definitions existed between studies.

Anti-nucleoprotein (NP) seropositivity, a marker of prior SARS-CoV-2 infection, was analyzed for the participants in these studies, and each study population showed a significant proportion of baseline seronegative individuals **(Table 1).** The four studies had similar median intervals between most recent pre-study and current study vaccinations (range 238 to 309 days, overlapping IQR) **(Table 1)**. Despite the use of samples from different clinical studies, post*-hoc* immunological testing of all samples was performed using the same assay methodologies.

### Correlation of anti-rS IgG subclasses with nAb and antibody-dependent Fc effector responses

The relationships between anti-S IgG subclass levels, nAb titers, and surrogates for antibody-dependent Fc effector functions were analyzed across a combined sample set of COVID-19 vaccinees who received four mRNA and/or protein-based rS doses of ancestral strain vaccines (2019nCoV-301 and 2019nCoV-307, N=61–74) **(Table 2)**. As there was not much difference in the IgG2 subclass between mRNA- and rS-primed groups, correlation analysis was not performed for anti-Spike IgG2. Spearman’s rank correlation analysis indicated a significant positive correlation between SARS-CoV-2 vaccine–induced anti-S IgG subclass levels with nAb (MN_50_) geometric mean titer (GMT) for IgG1 (0.83, 95% CI: 0.74 to 0.89, *P*<0.0001) and IgG3 (0.48, 95% CI: 0.28 to 0.64, *P*<0.0001) **(Table 2)**. By contrast, a significant negative correlation between IgG4 subclass with nAb GMT was observed (–0.39, 95% CI: –0.59 to –0.15, *P*=0.001) **(Table 2).**

**Table 2.** Correlations between IgG isotypes, neutralizing antibodies (MN50), and antibody-dependent Fc effector functions.

| Response Type | Assay | Anti-S IgG1<br>(N=72 to 74) | Anti-S IgG3<br>(N=73) | Anti-S IgG4<br>(N=61 to 63) |
| --- | --- | --- | --- | --- |
|  |  | Spearman's r<br>(95% CI)<br>*P* value | Spearman's r<br>(95% CI)<br>*P* value | Spearman's r<br>(95% CI)<br>*P* value |
| Neutralization | Microneutralization<br>(MN50) | **0.832**<br>(0.74 to 0.89)<br>*P*<0.0001 | **0.480**<br>(0.28 to 0.64)<br>*P*<0.0001 | **-0.394**<br>(-0.59 to -0.15)<br>*P*=0.001 |
| Antibody-Dependent<br>Fc Effector Functions | ADCP<br>(surrogate<br>FcγRIIa binding) | **0.917**<br>(0.87 to 0.95)<br>*P*<0.0001 | **0.533**<br>(0.34 to 0.68)<br>*P*<0.0001 | **-0.404**<br>(-0.60 to -0.16)<br>*P*=0.001 |
|  | ADCC<br>(surrogate<br>FcγRIIIa binding) | **0.957**<br>(0.93 to 0.97)<br>*P*<0.0001 | **0.592**<br>(0.41 to 0.73)<br>*P*<0.0001 | **-0.532**<br>(-0.69 to -0.32)<br>*P*<0.0001 |
|  | ADCD<br>(C1q binding) | **0.922**<br>(0.88 to 0.95)<br>*P*<0.0001 | **0.609**<br>(0.43 to 0.74)<br>*P*<0.0001 | **-0.528**<br>(-0.69 to -0.31)<br>*P*<0.0001 |

Data below the lower limits of quantitation were excluded from the analysis.

Similar response trends were observed in surrogate markers for antibody-dependent Fc effector functions. Positive correlations were observed for IgG1 and IgG3 for antibody-dependent cellular phagocytosis (ADCP) (0.92, 95% CI: 0.87 to 0.95, *P*<0.0001; and 0.53, 95% CI: 0.34 to 0.68, *P*<0.0001); antibody-dependent cellular cytotoxicity (ADCC) (0.96, 95% CI: 0.93 to 0.97, *P*<0.0001; and 0.59, 95% CI: 0.41 to 0.73, *P*<0.0001), and antibody-dependent complement deposition (ADCD) (0.92, 95% CI: 0.88 to 0.95, *P*<0.0001; and 0.61, 95% CI: 0.43 to 0.74, *P*<0.0001), respectively **(Table 2)**. By contrast, IgG4 levels were negatively correlated with antibody-dependent Fc effector functions ADCP (–0.40, 95% CI: –0.60 to –0.16, *P*=0.001), ADCC (–0.53, 95% CI: –0.69 to –0.32, *P*<0.0001), and ADCD (–0.53, 95% CI: –0.69 to –0.31, *P*<0.0001) **(Table 2).**

### Vaccine platform**–**specific immune priming elicits differential anti-rS IgG subclass and anti-rS Fc-Mediated Effector responses

To determine the role of COVID-19 vaccine platform–specific priming, anti-S IgG subclass levels pre- (day 0) and post-vaccine (day 28/29) were measured following administration of different homologous and heterologous vaccine dosing regimens of the ancestral strain **(Table 1**, **Fig. 1A)**. All groups showed an increase in S-specific IgG1, IgG2, IgG3, and IgG4 subclass levels post-dose compared to pre-dose levels (fold rise 1.6X–39.2X) **(Fig. 1B–I)**. Compared to priming with an adjuvanted rS protein, priming with mRNA was associated with lower overall IgG1 and IgG3 levels (161,964, 95% CI 128,002 to 204,937 and 504.5, 95% CI 328.5 to 774.8 vs 22,605.4, 95% CI 16,033 to 31,871 and 34.2, 95% CI 20.7 to 56.4, respectively) and higher IgG4 levels (48.7.9, 95% CI 17.3 to 137 vs 8567, 95% CI 3808 to 19,273) after the ancestral strain booster dose in this analysis. There was no significant difference in IgG2 levels between mRNA- and rS-primed groups (**Fig.1C**) either at pre- or post-boost. However, there was a significant increase (P<0.0001) in IgG2 levels post-boost in rS prime followed by the mRNA booster group (Fig.1G). Among the four-dose regimens, IgG1 and IgG3 responses were significantly higher (*P* =0.0002 - <0.0001), while IgG4 was significantly lower (*P* <0.0001), among adjuvanted protein-primed participants compared to those who received mRNA priming doses **(Fig. 1B–E).** Due to differences in the total number of doses received, data and statistical analysis of the protein-based rS + BNT162b2 mRNA group is shown separately in **Fig. 1 F–I.**

**Fig. 1:**
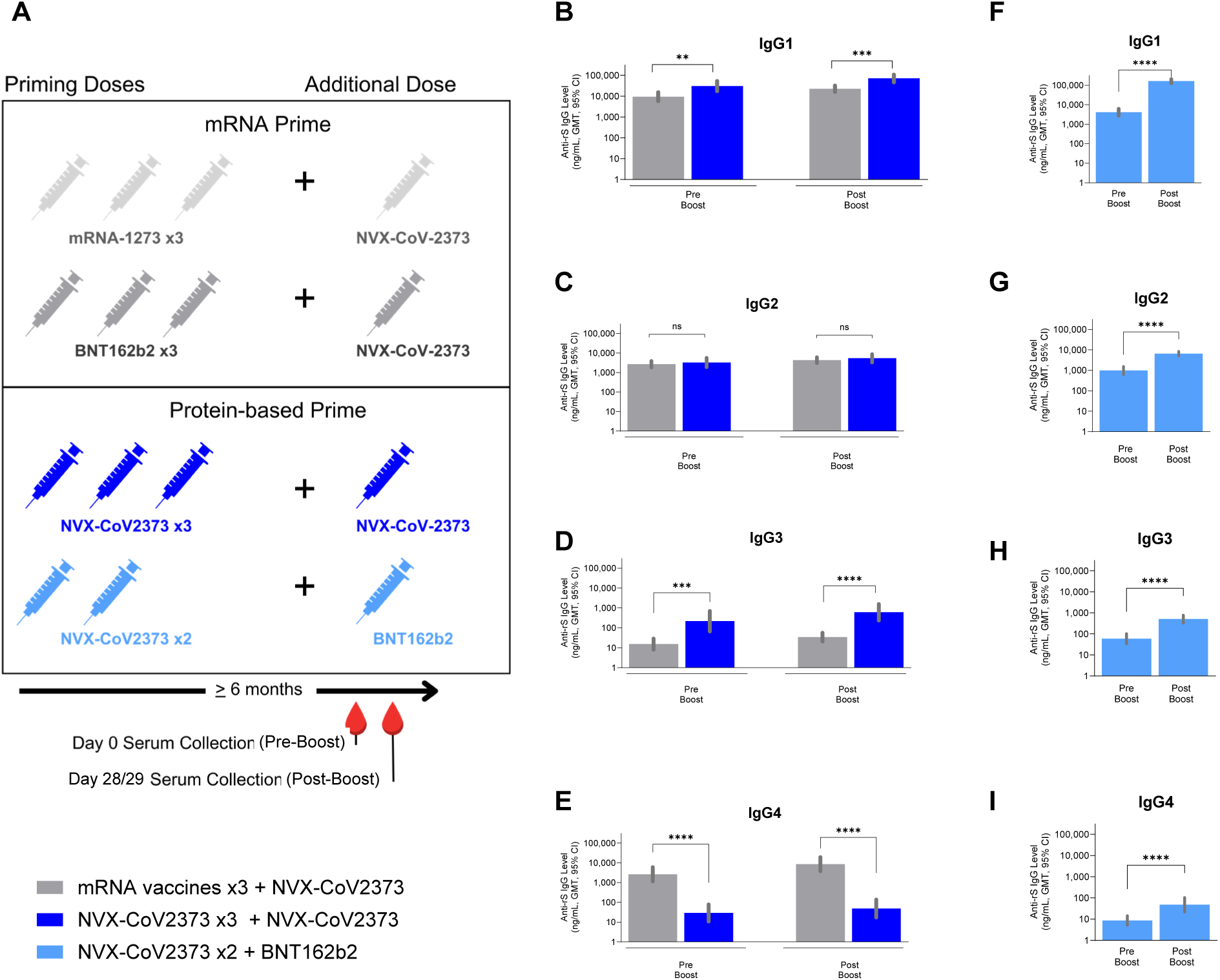
Anti-Spike (rS) IgG isotypes in subjects primed with either with mRNA vaccines (mRNA-1273 or BNT162b2) or the protein-based subunit COVID-19 vaccine (NVX-CoV2373). Pre-booster (day 0), post-booster NVX-CoV2373 (day 28) or mRNA (BNT162b2) vaccines (day 29). **A.** Schematic of the vaccines administered to subjects. ELISA-based quantitation [Geometric Mean Titers (GMT) with 95% confidence intervals (CI)] of IgG isotypes is shown in **B–E**: Gray – mRNA vaccines x3 +NVX-CoV2373 (N=19), dark-blue – NVX-CoV273 x3 + NVX-CoV273 (N=18), **F–I:** Light-blue – NVX-CoV273 x2 + BNT162b2 (N=24 for Day 0 and N=30 for Day 29). Statistical analysis was performed by the two-tailed Mann–Whittney test. **p<0.01, ***p < 0.001, ****p < 0.0001.n.s. indicates not significant.

For all IgG subclasses, the greatest fold-rise from pre-boost to post-boost was observed in participants who received the adjuvanted rS protein–based vaccine as two priming doses followed by an additional dose of BNT162b2 mRNA (Group C, **Table 1**) (IgG1, 39.2X; IgG2, 6.6X; IgG3, 8.6X; and IgG4 5.5X). The comparison of these fold increases is, however, confounded by the fact that this group (Group C, **Table 1**) received one fewer dose (three total doses) than the other comparator groups (four total doses, Groups A and B, **Table 1**). A statistically significant difference (*P* =0.0002 to <0.0001), in IgG1, IgG3, and IgG4 (but not IgG2) was observed between mRNA vaccine–-primed and adjuvanted rS-primed participants **(Fig. 1B–E)**. In participants who received two priming doses of rS followed by an additional dose of BNT162b2 mRNA (Group C, **Table 1**), a statistically significant (*P* <0.0001) increase in all IgG isotypes (IgG1–4) was observed at post-booster compared to pre-booster **(Fig.1 F–I)**; however, the IgG4 levels in this group (Group C, **Table 1**) were still lower than 50 ng/mL even at post-booster. Although pre- to post-booster-fold rises for IgG1-4 in recipients of two priming doses of rS followed by an additional dose of BNT162b2 mRNA were the highest, the overall IgG4 level was the lowest (47.9 ng/mL) of all regimen groups. Overall, IgG4 levels were notably higher (>150x) in participants primed by mRNA vaccines (8567.1, 95% CI 3808 to 19,273 ng/mL) compared with those primed by the rS protein–based vaccine (48.7, 95% CI 17.3 to 137.0 and 47.9, 95% CI 22.2 to 103.3 ng/mL for groups B and C, respectively). A similar trend was observed in participants who were either anti-NP (nucleoprotein)–positive (seropositive) **(Fig.s1a**) or –negative (seronegative) **(Fig.s1b).**

Irrespective of vaccine regimen, all anti-rS Fc effector responses were, as expected, higher post-additional dose than before the study vaccine (fold-rise 2.8X–38.6X) **(Fig. 2)**. Among recipients of four total doses, surrogate ADCP, ADCC, and ADCD responses were higher (*P* = 0.0039–0.0004) in rS-primed participants compared to those with mRNA priming **(Fig. 2A–C).** Due to differences in the total number of doses administered, data and statistical analysis of the NVX-CoV2373 + BNT162b2 group are shown separately in **Fig. 2 D–F**. A statistically significant difference (P<0.0001 to P=0.0039) in ADCP, ADCC, and ADCD was observed between participants with mRNA vaccine–primed and rS-primed subjects (**Fig. 2A–C**). In participants who received the rS protein–based vaccine as two priming doses followed by an additional dose of BNT162b2 mRNA (Group C), a statistically significant increase (*P* <0.0001) in Fc effector functions (ADCP, ADCC, and ADCD) was observed post booster compared to prior to booster (**Fig.2 D–F**). For all anti-S Fc effector responses, the greatest pre- to post-booster increase was again observed in participants primed with two doses of the rS protein–based vaccine and an additional dose of BNT162b2 mRNA (ADCP, 12.4X; ADCC, 7.9X; ADCD, 38.6X) **(Fig. 2).** Overall, the anti-rS Fc–mediated effector responses mirrored the immune response trends of IgG1 and IgG3, as the rS protein–based vaccine led to higher post-additional dose ADCP (MFI), ADCC (MFI), and ADCD (MFI) function scoring (ADCP= 11,355, 95% CI 7826 to 16,476 and 37,361, 95% CI 35,928 to 38,852; ADCC=14,878, 95% CI 10,859 to 20,386 and 46,489, 95% CI 44,854 to 48,182; ADCD= 36,532 95% CI 21,236 to 62,844 and 97,736, 95% CI 92,572 to 103,187 for groups B and C, respectively) than mRNA priming (Group A, GMT of ADCP= 5776, 95% CI 4268 to 7815; ADCC=5650, 95% CI 3748 to 8518; ADCD=6369, 95% CI 2705 to 14,994) **(Fig. 2).** A similar trend was observed in participants regardless of the pre-booster anti-NP serostatus **(Fig.s2a and b).**

**Fig. 2:**
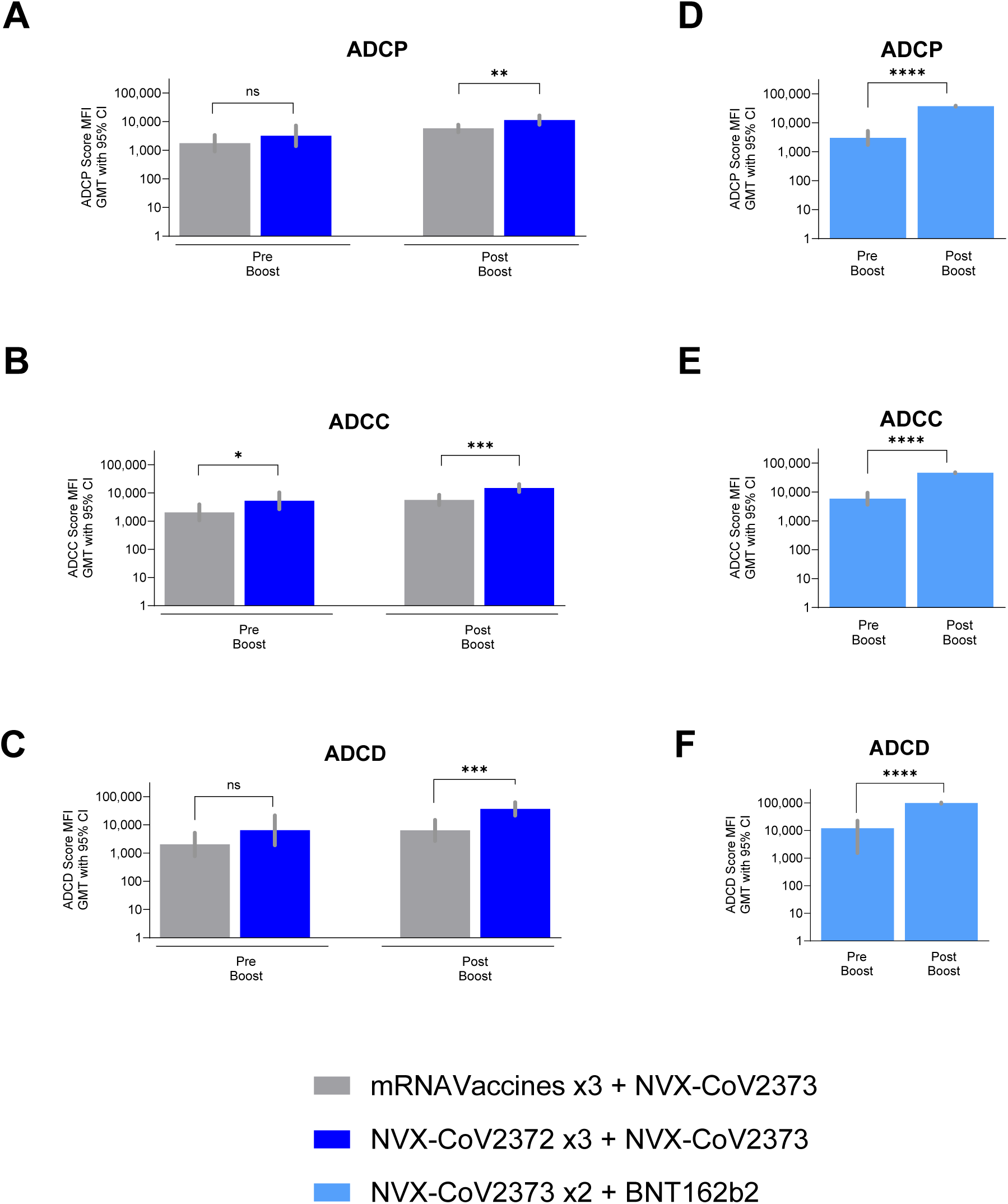
Fc-effector assay (surrogate assays for ADCP, ADCC, and ADCD) data in subjects primed either with mRNA vaccines (mRNA-1273 or BNT162b2) or the protein-based subunit COVID-19 vaccine (NVX-CoV2373). Pre-booster (day 0), post-booster NVX-CoV2373 (day 28) or mRNA (BNT162b2) vaccines (day 29). GMT with 95% CI is shown. **A, D:** Surrogate assays for ADCP (FcgR2a binding), **B, E:** Surrogate assays for ADCC (FcgR3a binding), **C, F:** Surrogate assays for ADCD (C1q binding). **A–C:** Gray – mRNA Vaccines x3 +NVX-CoV2373 (N=20), dark-blue – NVX-CoV273 x3 + NVX-CoV273 (N=17), D-: light-blue – NVX-CoV273 x2 + BNT162b2 (N=24 for Day 0 and N=30 for Day 29). Statistical analysis was performed by the two-tailed Mann–Whittney test. p<0.05, **p < 0.01, ***p < 0.001, indicates “significance”. n.s. indicates not significant.

### Strain-specific IgG1 and IgG4 immune imprinting following previous mRNA prime

The impact of vaccination with updated COVID-19 formulations based on newer SARS-CoV-2 variants was assessed by comparing IgG responses specific for the rS ancestral strain to those based on XBB.1.5 rS protein. Here, all participants received mRNA prime (≥3 prior ancestral mRNA doses) followed by an additional adjuvanted rS dose with updated XBB.1.5 strain–based rS (NVX-CoV2601, study 2019nCoV-313, Group D, **Table 1**). IgG1 and IgG4 comprised 98% or more of total anti-S IgG both pre- and post-additional doses **(Fig. 3A and B).** The anti-ancestral rS and anti-XBB.1.5 rS proportions of IgG1/IgG4 were similar pre- and post-additional dose, though some differences between rS strains were observed **(Fig. 3A and B)**. Anti-ancestral rS responses elicited lower IgG1/IgG4 ratios compared to those induced by XBB.1.5 rS, suggesting potential immune imprinting in favor of ancestral rS following previous mRNA prime **(Fig. 3A and B)**. A similar trend was observed regardless of the anti-NP status **(Fig.s3a and b)**. Overall, in this small sample, mRNA1273 prime led to a lower IgG1/IgG4 ratio compared to BNT162b2 prime **(Fig. 3A and B)**.

**Fig. 3:**
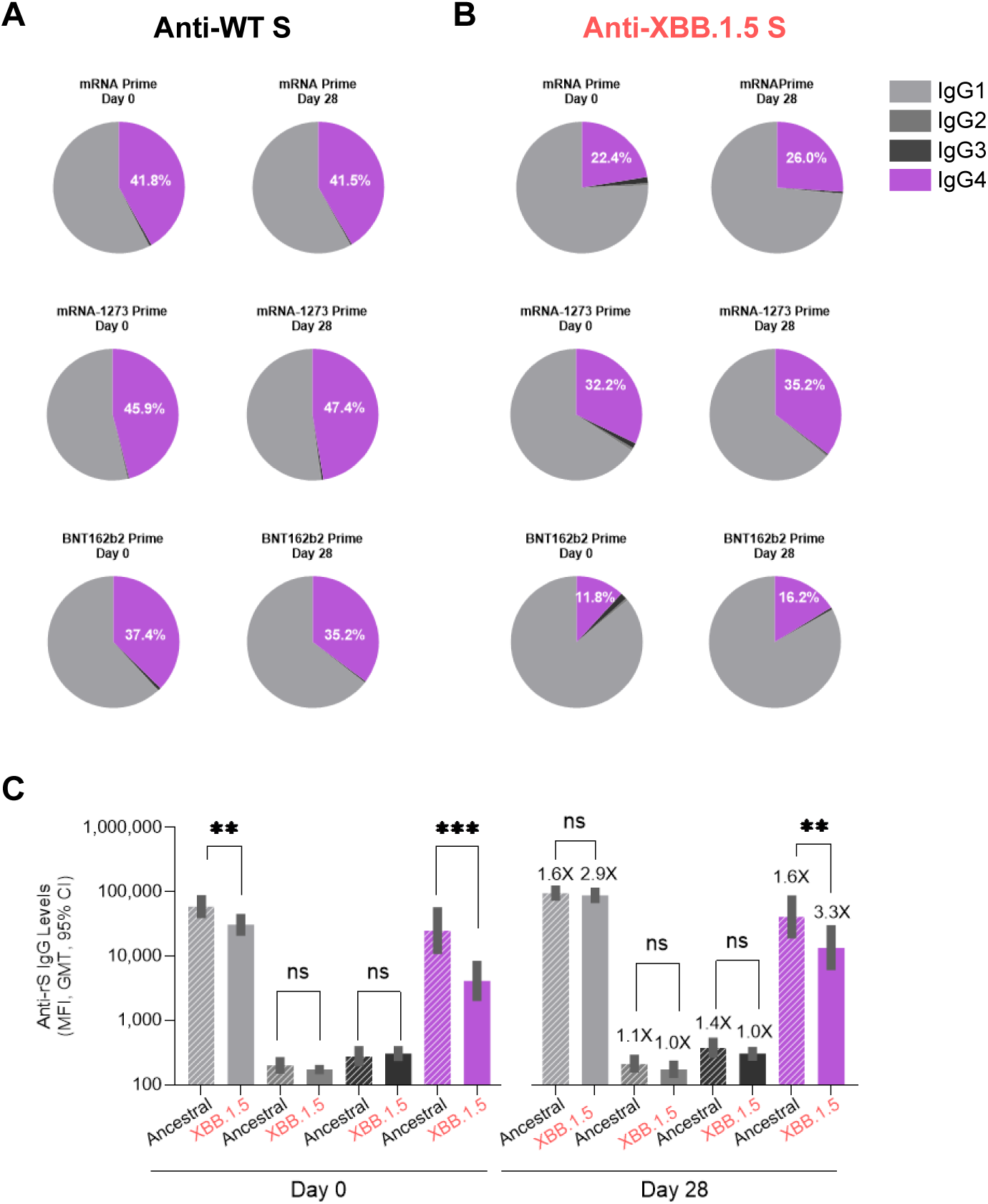
Anti-Spike (rS) IgG isotype proportions in subjects receiving the protein-based subunit XBB.1.5 COVID-19 vaccine (NVX-CoV2601) after priming with mRNA vaccines (≥3x). Pre-booster (day 0), post-booster with the NVX-CoV2601 vaccine (day 28). **A.** Anti-rS IgG isotype proportions (pie chart) against the wild-type (WT) trimer antigen (ancestral) **B.** Anti-rS IgG isotype proportions (pie chart) against the XBB.1.5 trimer antigen **C.** Anti-rS IgG isotype levels (GMT with 95% CI bar graph) against WT (Stripes, N=29) and XBB.1.5 (Solid color, N=29) trimer antigens. Geometric mean fold raise (GMFR) at day 28 post booster compared to pre-booster levels (day 0) are shown above the bars. Light-gray – anti-rS IgG1, medium-gray – anti-rS IgG2, black – anti-rS IgG3, plum – anti rS IgG4. Statistical analysis was performed by the two-tailed Mann–Whittney test. **p < 0.01, ***p<0.001 indicates significance. n.s. indicates not significant.

At Day 0, prior to receiving an additional dose with XBB.1.5 rS, there were low levels of IgG1 and IgG4 for XBB.1.5 (GMT=IgG1, 30,714, 95% CI 20,773 to 45,412; IgG4, 4119, 95% CI 1998 to 8491), although levels specific to ancestral rS were, unsurprisingly, higher (GMT=IgG1, 58,899, 95% CI 38,849 to 89,298; IgG4, 25,051, 95% CI 10,845 to 57,868) **(Fig. 3C).** Significantly higher ancestral-specific IgG4 compared to lower XBB.1.5–specific IgG4 at both pre- (GMT=25,051 vs. 4119) and post-additional (GMT=40,945, 95% CI 19,002 to 88,225 vs. 13,562, 95% CI 6079 to 30,253) dose likely reflects ancestral rS protein–specific immune imprinting from prior ancestral-based mRNA vaccination **(Fig. 3C)**. Fold-rises following the additional dose of XBB.1.5 rS were low to moderate for IgG1 and IgG4 cross-reactive with the ancestral strain (1.6X and 1.6X, respectively, and only slightly higher for XBB.1.5–specific IgG1 and IgG4 2.9X and 3.3X, respectively. Interestingly, the post-booster IgG1 levels to ancestral and XBB.1.5 strains were not significantly different (ns), despite their difference prior to the XBB.1.5 rS boost (*P* <0.01). This indicates the ability of the XBB.1.5 vaccine boost to at least partially overcome imprinting and induce XBB-specific antibodies in addition to ancestral-specific recall antibodies. By contrast, IgG2 and IgG3 levels were similar between anti-XBB.1.5 and anti-ancestral rS levels with overlapping 95% CIs (**Fig. 3C**). Overall, post-final booster dose levels of anti-XBB.1.5 rS IgG1 and IgG4 were higher than pre-booster (day 0) levels (GMT= 87,937 and 13,562 vs 30,714 and 4,119, respectively) and the difference between the anti-ancestral and XBB.1.5 seemed to be decreasing (fold decrease for ancestral/XBB.1.5 IgG1 from 1.9 to 1.1 and IgG4 from 6.1 to 3.0) **(Fig. 3C).** This reduction in the difference suggests that the original “antigenic sin” (immune imprinting) is being overcome by an XBB.1.5 strain–specific booster.

As RBD antigens between ancestral and XBB.1.5 strains show significant differences, IgG levels against RBD antigens were also measured. In contrast to the IgG levels against rS spike trimer antigens, the IgG levels against RBD antigens showed a stark, statistically significant (*P* <0.001 - <0.0001) contrast between ancestral and XBB.1.5 strains **(Fig.s3c_C; s3d-e)**, consistent with the presence of conserved epitopes in the Spike trimer, but not in RBD antigens. IgG1 proportions at baseline were higher (*P* <0.01) in seropositive subjects (GMT 89,625, 95% CI 65,084 to 123,419 and mean 68.75 %, SEM 5.9%) (**Fig s3a**, anti-NP^+^) than seronegative subjects (GMT 26,527, 95% CI 10,260 to 68,589 and mean 36.16% SEM 9.9%) **Fig s3b**, anti-NP^-^). Similarly, IgG4 levels and proportions were different between the two groups (GMT 17,283, 95% CI 5119 to 58,346 and mean 30.9 % SEM 5.9% for seropositive anti-NP+ (**Fig s3a)** vs GMT 50,716, 95% CI 21,214 to 121,249 and mean 62.6% SEM 9.9% for seronegative (**Fig.s3b)**.

### Strain-specific nAb and Fc effector responses reflect immune imprinting following previous mRNA prime

The samples from participants who received ancestral mRNA prime (three prior mRNA doses) followed by an additional dose with the updated XBB.1.5 strain–based rS protein–based vaccine (NVX-CoV2601) were analyzed. Pre-dose ancestral strain nAb titers (PNT based) (GMT=1392, 95% CI 851 to 2277) were higher than the XBB.1.5 nAb titers (GMT=141.1, 95% CI 68.4 to 291.0) **(Fig. 4A).** Ancestral strain nAb titers (detected in PNT) rose 1.9-fold and remained significantly higher (GMT=2640, 95% CI 1747 to 3989) (*P<0.05*) than XBB.1.5 nAb titers (GMT=1248, 95% CI 715–2180) after XBB.1.5 boost **(Fig.4A**) as a product of imprinting due to prior ancestral exposures and shared epitopes. Nonetheless, XBB.1.5 specific titers rose much more sharply, 8.8-fold indicating the induction of nAb recognizing new XBB.1.4 specific epitopes. Interestingly, the difference between nAb titers against ancestral and XBB.1.5 strains was lower after booster compared to pre-booster (ratio of ancestral/XBB.1.5, 2.12 vs 9.87 post-booster vs pre-booster, respectively), suggesting a possible effect of XBB.1.5 booster in partially overcoming the immune imprinting, which is similar to the trend observed in IgG subclass data. ADCP assay (FcgR2a binding), also suggested evidence of potential immune imprinting as post-additional dose, ADCP for the ancestral strain was notably higher (GMT=14,326, 95% CI 11326 to 18,120 vs. 8808, 95% CI 5964 to 13,007; P<0.05 **Fig.4B**) than the XBB.1.5 strain. By contrast, the ADCC assay (FcgR3a binding), and the ADCD assay (C1q Binding), did not show a statistical difference between ancestral and XBB1.5 strains (GMT=14,912, 95% CI 11,440 to 19,439, and 14,814, 95% CI 7579 to 28,955, respectively, for the ancestral strain and GMT=10,528, 95% CI 7278 to 15,230, and 9238, 95% CI 4992 to 17,094 for XBB.1.5 rS) **(Fig. 4B–D)**. Though overall ancestral strain responses were generally higher, presumably due to previous vaccinations and natural infection, a greater fold-rise in XBB.1.5 rS-specific responses induced by booster were seen (ADCP=5.1X; ADCC=4.4X; ADCD=4.8X) compared to only a slight increase in ancestral strain nAb (ADCP=1.7X; ADCC=1.7X; ADCD=2.1X) **(Fig. 4B–D)**. ADCP, ADCC, and ADCD levels did not show a difference between ancestral and XBB.1.5 strains regardless of the anti-NP status of subjects (**Fig.s4a–A, B, C, D**) and **Fig.s4b_A, B, C, D**), which might be due to the variant-agnostic attribute of the Fc effector functions. Analysis of nAb, ADCP, ADCC, and ADCD levels in anti-NP–negative subjects showed a similar trend, except nAb levels [which showed a significant (*P* <0.05) response to an XBB.1.5 booster].

**Fig. 4:**
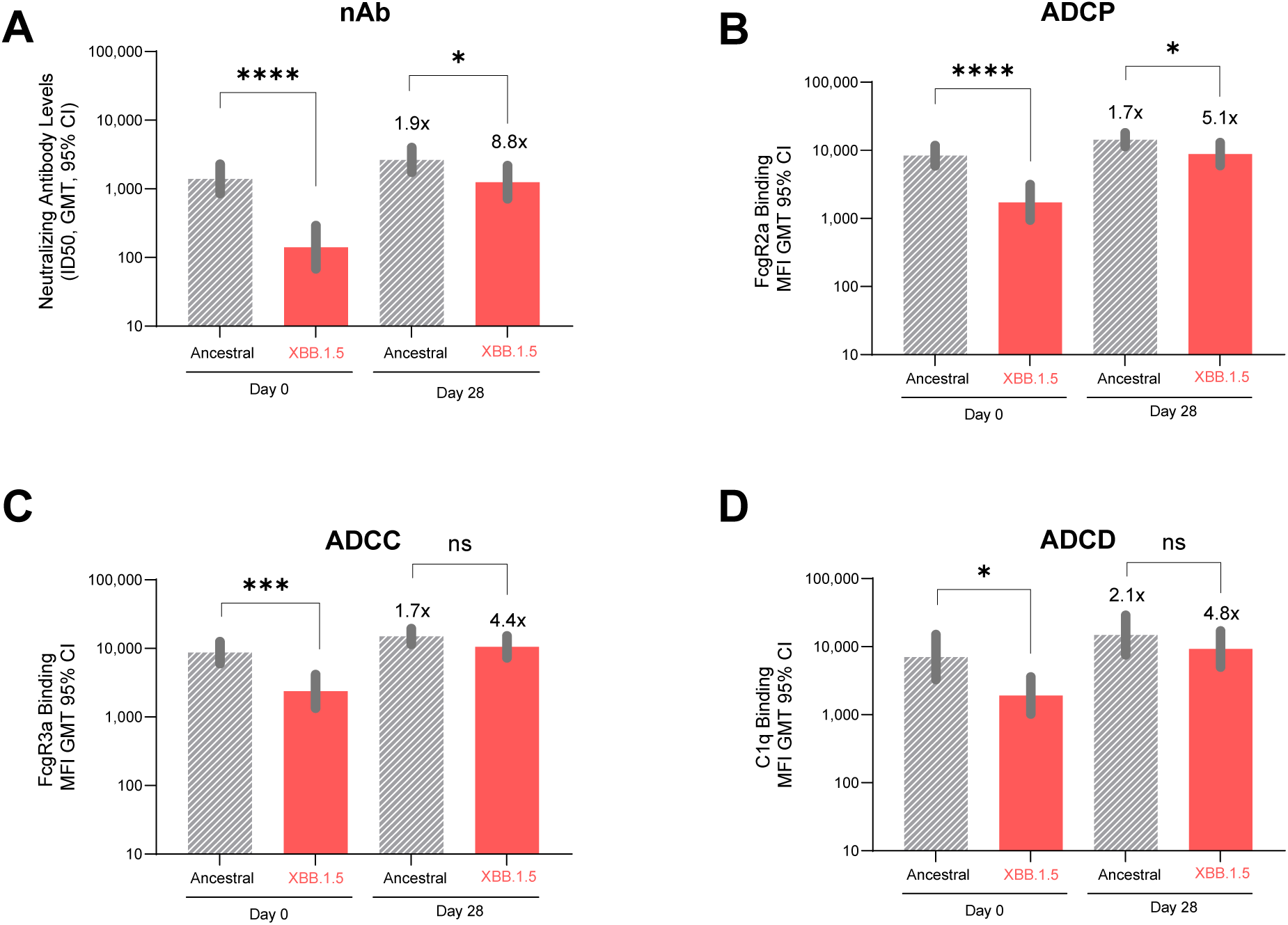
Neutralizing antibody levels (ID50) and Fc effector assay data (surrogate assays for ADCP, ADCC, ADCD) in subjects receiving the protein-based subunit XBB.1.5 COVID-19 vaccine (NVX-CoV2601) after priming with mRNA vaccines (≥3x). Pre-booster (day 0), post-booster with the NVX-CoV2601 vaccine (day 28). **A**. Neutralizing antibody titers against ancestral (WT) (N=29) and XBB.1.5 (N=29) strains measured by pseudovirus neutralization assay **B.** Surrogate assays for ADCP (FcgR2a binding), against ancestral (WT) (N=29) and XBB.1.5 (N=29) trimer antigens **C.** Surrogate assays for ADCC (FcgR3a binding), against ancestral (WT) (N=29) and XBB.1.5 (N=29) trimer antigens. **D.** C1q binding for ADCD, against ancestral (WT) (N=29) and XBB.1.5 (N=29) trimer antigens. GMT with 95% CI is shown. Geometric mean fold raise (GMFR) at day 28 post booster compared to prebooster levels (day 0) are shown above the bars. Gray stripes – ancestral, red – XBB.1.5. Statistical analysis was performed by the two-tailed Mann–Whittney test. *p < 0.05,***p<0.001, ****p<0.0001 indicate significance. n.s. indicates “not significant”.

Overall, the observed increase in XBB.1.5 rS-specific responses was somewhat greater for functional immune responses (nAb and Fc effector functions) than IgG1 and IgG4 following the XBB.1.5 rS protein-based vaccine (and may suggest a partial overcoming of immune imprinting for functional immunity specific for ancestral S that could be beneficial) **(Fig. 3 and 4; Fig. s4d–e).**

## Discussion

This is the first evidence of a negative correlation between increased SARS-CoV-2 vaccine–induced anti-S IgG4 levels with neutralizing antibody titers, which might be due to the Fab arm exchange described in the literature for IgG4^2^. In agreement with previous research^8^, we find evidence that SARS-CoV-2 vaccine–induced anti-S IgG4 levels are also inversely correlated with surrogate markers for Fc-effector functions that may contribute to the COVID-19 immune response. The long-term clinical implications of vaccine-mediated increases in anti-S IgG4 requires additional evaluation. In parallel, we show differential immune priming of specific antibody-dependent Fc effector responses associated with specific COVID-19 vaccine platforms. The distinctive characteristics of the vaccine immunogens produced by different platforms (adjuvanted recombinant S protein vs. mRNA coded, host-produced S protein), or the differences in their presentation to T cells *in-vivo*, that are responsible for the observed unique differences remain uncertain and warrant future investigation.

Further, we demonstrate that immune priming induced by the first 2–3 vaccine doses appear to influence the development of a unique anti–S-specific IgG4 response. Elevated IgG4 levels and proportional increases in IgG4 as a percentage of total IgG were observed in those who were primed with mRNA COVID-19 vaccine. By contrast, priming with the adjuvanted rS protein–based vaccine followed by additional doses with either rS or mRNA vaccine yielded a comparatively negligible increase in anti-S IgG4 that was >150X lower than that of participants primed by mRNA vaccines. The potential functional impact of this phenomenon was observed as mRNA priming was associated with lower nAb titers and reduced Fc effector responses (ADCP, ADCC, and ADCD) compared to adjuvanted rS priming. We also observed higher IgG1 proportions at baseline in seropositive subjects **(Fig.s3a)** than seronegative subjects (**Fig s3b**, anti-NP^-^), which might be due to prior infections.^11^ IgG4 levels and proportions were vice versa between the two groups (**Fig s3a vs Fig.s3b**), which might be due to prior infection influencing IgG4 switch.^11^

This study was limited due to the use of relatively small numbers of clinical samples from several trials. This resulted in potential differences in cohort characteristics, timing of sample collection, unknown variations in participant histories of natural infection, and small sample sizes per study. Thus, comparisons of antibody levels between groups may not represent the broader population. In future studies, the addition of baseline control groups (i.e., no vaccine, placebo, or convalescent serum) would aid in the interpretation of the relevance of the observed immune responses. Notably, this study does not address long-term protective vaccine effectiveness and safety signals essential for understanding overall benefits in real-world settings.

Several hypotheses associated with proportional increases in vaccine-antigen specific IgG4 to infectious disease should be considered. IgG4 exhibits a reduced capacity to activate Fc effector responses, such as ADCC, ADCP, and ADCD. ^29^ Compared to nAbs, Fc effector functions have been shown to be more broadly cross-reactive to antigenically diverse SARS-CoV-2 variants^14,30–32^. We have also observed a correlation to lower levels of nAbs, which are considered an immune correlate of protection against COVID-19^33–35^. Here, we hypothesize that while mRNA priming with ancestral strain–based vaccines may generate persistent immune imprinting that biases the immune response to the production of anti-ancestral rS protein–specific IgG4, administration of an updated protein-based COVID-19 vaccine such as XBB.1.5 (or newer variants such as JN.1) may avoid exacerbation of the IgG4 class switch. In this study, ≥3-dose ancestral mRNA priming followed by an additional dose of the updated XBB.1.5 rS protein–based vaccine led to moderate increases from baseline anti-S IgG4 but also led to marked increases in XBB.1.5–specific nAb and Fc effector functions, although these increases were lower in magnitude than were induced by the ancestral vaccine.

Most vaccine recipients in the United States (US) were administered initial vaccination series with one or more additional doses with mRNA vaccines. An intriguing future direction of this work could include the investigation regarding what, if any, future influence the general population’s immunological background may have on the quality of immune responses to subsequent COVID-19 vaccination. According to several recent studies^8^ and our own findings, 2–3 doses of ancestral mRNA COVID-19 vaccination led to increased production of anti-ancestral S IgG4, while an adjuvanted rS protein–based vaccine regimen did not. While ancestral strain imprinting, which apparently includes an mRNA-induced bias toward IgG4 responses to ancestral epitopes, may not be reversible, it remains to be determined if additional vaccination with an antigenically distinct updated formulation of the COVID-19 rS protein-based vaccine (such as the XBB.1.5–based vaccine [(NVX-CoV2601] used in this study or future newer variants such as JN.1) could avoid exacerbation of the observed anti-S IgG4 response and potentially take advantage of naive responses to the distinctive epitopes of newly emerged strains. A recent publication^36^ showed that after an XBB.1.5 mRNA vaccine booster, immune imprinting was still observed, and the immune response was dominated by B-cell memory recall response.

As the clinical impact of IgG subclasses and Fc effector functions in the context of SARS-CoV-2 infection remains unknown, their relative contribution along with other aspects of immunity (i.e., T-cell activity and mucosal immunity) must be studied further. There remains uncertainty regarding immune tolerance to subsequent additional doses and the impact of antigenic changes in vaccine antigens and circulating SARS-CoV-2 strains.

## Methods

### Study participants and clinical studies

Human serum samples were obtained from clinical trials 2019nCoV-301 (Novavax), 2019nCoV-307 (Novavax), 2019nCoV-313 (Novavax), and VRC433 (University of Washington). Relevant consent forms included exploratory assays of SARS-CoV-2 antibody response measurements. Study demographics are shown in **Table. 1**. The 2019nCoV-301 (PREVENT-19) study was a phase 3, randomized, observer-blinded, placebo-controlled trial in the US and Mexico during the first half of 2021 to evaluate the protective efficacy and safety of NVX-CoV2373 in adults (≥18 years of age). Additional study details can be found at clinicaltrials.gov, NCT04611802^37^. The 2019nCoV-307 study was a phase 3, trial evaluating immunogenicity and safety of NVX-CoV2373 to confirm lot-to-lot consistency in previously vaccinated adults aged 18–49 years in the US. Additional study details can be found at clinicaltrials.gov, NCT05463068^38^. The 2019nCoV-313 (part 1) study was a phase 2/3 open-label, single-arm study of NVX-CoV2601 in participants who were aged ≥18 years and previously vaccinated with ≥3 doses of mRNA-1273 or BNT162b2. Co-primary objectives of this 28-day immunogenicity and safety assessment were to determine if one intramuscular injection of NVX-CoV2601 (5 µg XBB.1.5 S + 50 µg Matrix M™ adjuvant) induced superior neutralizing antibody responses to XBB.1.5 compared to a historical control group (2019nCoV-311/NCT05372588 [part 2] study participants) who received ancestral strain adjuvanted rS (NVX-CoV2373) after three prior mRNA-based vaccinations. Additional study details can be found at clinicaltrials.gov, NCT05975060. In each clinical trial, serum samples to be tested for immune responses were obtained from participants at baseline prior to the vaccine dose of interest and at least 14–29 days after that dose. Participants were also followed for safety for 1–6 months after the vaccine dose of interest. VRC433 at the University of Washington was an observational study of participants previously enrolled in the PREVENT-19 study (2019nCoV-301) to evaluate the immune response of persons who had received priming doses of NVX-CoV2373 at 21 days apart (clinicaltrials.gov, NCT04611802). These participants received a dose of a commercial mRNA vaccine (BNT162b2 (30- μg mRNA)) and continued in safety follow-up for ≥6 months after the priming doses of NVX-CoV2373.

### IgG Subclass Measurements

The anti-S IgG4, IgG1, and total IgG assays were developed to measure the levels of anti-S IgG and % of Total anti-S IgG in serum samples from subjects immunized with COVID-19 vaccines or COVID-19 convalescent subjects. Anti-S IgG1, IgG2, IgG3, and IgG4 ELISA assays were performed as fit-for-purpose at Novavax Clinical Immunology lab (Gaithersburg, MD) using the previously published method^40^, with slight modifications. An anti–SARS-CoV-2 S RBD IgG1/IgG2/IgG3/IgG4 monoclonal antibody (Acro Biosystems, Newark, DE, USA) were used as reference standards for each respective assay. Assay plates (Thermo Fisher Scientific, Waltham, MA, USA) were coated with 1–2 µg/mL SARS-CoV-2 rS protein (produced at Novavax, Inc., Gaithersburg, MD, USA) overnight at 2–8 °C. Plates were then washed with phosphate-buffered saline with Tween 20 (PBST) and blocked for 1 h with blocking buffer (Thermo Fisher Scientific). Human serum samples together with isotype matched reference standards were then added to the wells, allowing anti-rS protein IgG antibodies to bind to S protein coated on plate (2 h of incubation). The plates were again washed with PBST, and then a secondary antibody: mouse anti-human IgG1 (Sino Biological), a mouse anti-human IgG2 (SouthernBiotech), a mouse anti-human IgG3, (Sino Biological) a mouse anti-human IgG4 (Invitrogen) conjugated with horseradish peroxidase was added and incubated for 1 h at room temperature (RT). A final wash was performed, followed by addition of 3,3’5,5’-tetramethylbenzidine substrate (TMB, Thermo Fisher Scientific). The reaction was stopped after 25 min by TMB stop solution (Scytek Laboratories, Logan, UT, USA). The optical density (OD) of the chromogenic signal proportional to the amount of anti-rS IgG captured on the plate, provided a quantifiable measurement of IgG isotype concentration in the serum sample. Anti-S IgG1/IgG2/IgG3/IgG4 concentrations were calculated by interpolating to the levels of the IgG reference standard curve. Data for anti-S IgG1, IgG2, IgG3, and IgG4, for individual subjects were tabulated. Anti-S IgG isotypes (%) were calculated by the ratio of isotype over total anti-S IgG*100 for each subject.

#### Microneutralization Assay

Microneutralization (MN) assay to measure NAbs was performed by the validated live virus method at 360biolabs (Melbourne, Australia) described previously^41^. Briefly, heat-inactivated human sera were diluted in medium comprised of Dulbecco Essential Medium without *L*-glutamine (DMEM; Thermo Fisher Scientific, Cat. No. 10313–021) supplemented with 2% fetal bovine serum (FBS; Bovagen SFBS), 1% GlutaMAX™ (Thermo Fisher Scientific, Cat. No. 35050–061), and 1% pen/strep (Thermo Fisher Scientific, Cat. No.15140–122), in an 11-point, 2-fold serial were mixed with an equal volume (140 μL) of SARS-CoV-2 virus (4000 TCID50 units/mL for the ancestral or XBB.1.5 strains and incubated for 1 h at 37 °C, 5% CO_2_. Following this incubation, 100 μL of the virus/serum mixtures (200 TCID50 units/well for the ancestral strain) were added in duplicate to Vero E6 cells, preseeded 24 h prior in 96-well plates in 100 μL of assay media at 1.5 ×10^4^ cells/well. Plates were incubated for 3 days at 37 °C, 5% CO_2_. The residual non-neutralized virus was detected via cytopathic effect (CPE) assessed by microscopic scoring. The neutralization titer was expressed as the reciprocal of the highest dilution at which ≥50% of the replicate wells were protected from infection (MN_50_).

### Surrogate Fc effector function multiplex assays

FcγRIIa and FcγRIIIa dimer binding via a multiplex assay were used as a surrogate for ADCP and ADCC, respectively, performed at the Chung Immunology Lab (University of Melbourne, Vic, Australia) using fit-for-purpose methods described previously^42^. Briefly, a customized cocktail of SARS-CoV-2 antigensand influenza HA (positive control) (proteins all from SinoBiological), were coupled to magnetic carboxylated multiplex beads (BioRad) as previously described^42^. Twenty microliters of working bead mixture containing 1000 beads per bead region and 20 μL of diluted sera (1:3200 dilution) were added to each well of a black, clear-bottom 384-well plate (Greiner Bio-One). Serum dilutions were selected by identifying the dilution that best correlated with serum ED50s as previously described^43^. After an overnight incubation at 4 °C on a shaker, plates were washed with PBS containing 0.05% Tween-20 (PBST). Soluble biotin labelled dimerized recombinant FcγR dimers (higher affinity polymorphisms FcγRIIa-H131 and FcγRIIIa-V158) were added to the wells and further incubated at RT for 2 h, followed by wash and addition of streptavidin, R-Phycoerythrin conjugate (SAPE, Invitrogen) at 1ug/mL, 25 uL/well. Plates were incubated at RT for 2 h on a shaker followed by washing and reading on a FlexMap 3D. Binding of PE detectors was measured to calculate the median fluorescence intensity (MFI). For the detection of C1q, biotinylated and tetramerized C1q protein (MP Biomedicals) with SAPE were added at 1 μg/mL, 25 μL per well, incubated at RT for 2 h on a shaker and then washed, followed by reading by the Luminex® FLEXMAP 3D. Binding of PE detectors was measured to calculate the median fluorescence intensity (MFI) followed by double background subtraction (removal of blank background and SIV gp120 negative control background). Assays were repeated in duplicate.

### Multiplex-based IgG subclassing

The multiplex-based IgG subclass assay was performed at the Chung Immunology Lab (University of Melbourne, Vic, Australia) using fit-for-purpose methods described by Brown et al and Selva et al.^42,44^ The same customized cocktail of SARS-CoV-2 multiplex beads as described above (1000 beads per bead region) along with 20 μL of diluted serum (1:1600 working dilution) were added to a black clear-bottom 384-well plate (Greiner Bio-One) followed by incubation overnight at 4 °C on a shaker and washing with PBS containing 0.05% Tween-20 (PBST). Strain-specific antibodies were detected using biotin-conjugated mouse anti-human IgG1–4 at 1.3 μg/mL, 25 μL per well for 2 h on a shaker, followed by wash and the addition of streptavidin PE at 1 μg/mL, 25 μL per well. The plates were washed after incubation at RT for 2 h on a shaker, followed by resuspension in 50 μL of sheath fluid, and reading by FlexMap3D. Median fluorescence intensity of PE bound beads was calculated, followed by double background subtraction (removal of blank background and SIV gp120 negative control bead background).

### Anti-NP assays

Anti-NP antibody detection assays were conducted at the University of Washington using the validated Roche Elecsys Anti-SARS-CoV-2 electrochemiluminescence immunoassay (ECLIA) assay that detects the nucleocapsid (N) antigen (Roche-N), and Roche cobas® e 411 analyzer as per the manufacturer’s kit protocol.

### Pseudovirus neutralization assay

The pseudovirus neutralization assay was performed following validated method^44^ in human samples. Briefly, serum samples were heat-inactivated in a 56°C water bath for 30 minutes prior to assay. Serum samples were serially diluted three-fold in duplicates in infection medium (DMEM without phenol red + 2%FBS + Penicillin + streptomycin + glutamine) at 1:10 starting dilution which was reported as 1:20 after addition of virus in a 96-well white opaque cell culture plate. Fifty microliters of working dilution Pseudovirus (corresponding to 50,000 RLU) was then added to each well, followed by incubation at 37°C for two hours. Then, 1.0 × 10^4^ HEK293T/hACE2 cells (obtained from Creative Biogene) in 100 µL of infection medium were added to the wells, followed by incubation for 72 hours at 37°C. After incubation, 50 µL BrightGlo Luciferase Substrate (Promega) was added to each well. Plates were incubated for 15 minutes at room temperature without ambient light. Viral entry into the cells was determined by measuring the luminescence with a SpectraMax iD3 microplate reader. Neutralization titers represent the inhibitory dilution of serum samples at which RLUs were reduced by 50% compared to virus control wells on each plate. Data were analyzed and inverse serum dilutions at which 50% pseudovirus Neutralization was observed (ID_50_) were calculated using 4-parameter curve fitting in SoftMax Pro software.

### Statistical analysis

Linear regression analyses were conducted using GraphPad Prism® software (San Diego, CA, USA; Version 10.2.1) and SAS® software (v9.4, SAS Institute Inc., Cary, NC, USA)

## Data Availability

All data produced in the present work are contained in the manuscript.

## Acknowledgments

The sponsor acknowledges the contributions and participation of all 2019nCoV-301, 2019nCoV-307, and 2019nCoV-313 study volunteers, principal investigators, and investigative site personnel who contributed to the success of the studies. Help from the University of Washington in performing the anti-NP analysis and 360biolabs in Melbourne, Vic, Australia for determining Nab is much appreciated. We thank Bruce D. Wines and P. Mark Hogarth, the Burnet Institute, Melbourne, Vic, Australia, for sharing their Fcγ Receptor dimer plasmids. The authors thank Benjamin Haner and team for their contribution to the pseudovirus neutralization assay. The authors also thank Anar Murphy, PhD, of Novavax, Inc. for editorial support.

## Author Contributions

Conceptualization: R.K., M.Z., A.W.C., S.C.C., J.S.P., A.M.M., L.M.D., and L.F.

Investigation: R.K., M.Z., A.W.C., S.C., J.S.P., A.P., D.G., M.R.C., S.M., P.R., L.C.A., T.M.B., A.W., and K.J.S.

Data Analysis: R.K., M.Z., A.W.C., S.C., A.P., D.G., Z.C., G.S., M.R.C., S.M., P.R., and J.S.P.

Visualization: R.K., and A.M.M.

Writing - original draft: A.M.M.

Writing - review and editing: All authors.

Project Administration: R.K.

## Declaration of interest statement

The authors declare the following financial interests/personal relationships which may be considered as potential competing interests: R.K., M.Z., S.C.C., A.P., D.G., Z.C., M.R.C., S.M., G.C., A.M.M., and J.S.P. are current or former employees of Novavax, Inc. and as such receive a salary and may hold Novavax, Inc. stock. L.F. and L.M.D. are consultants to Novavax, Inc. A.W.C. has received grant funding from NHMRC, MRFF, and NIH. A.W, received research funding from NIH, GSK, Assembly Biomedical and Moderna, and is a consultant for GSK, Aicuris, Innovative Molecules and Bayer. T.M.B. has served on an advisory board for Sanofi. P.R., L.C.A., and K.J.S. have no competing interests to declare.

## Notes

### Clinical Trial

NCT04611802; NCT05463068; NCT05372588; NCT05975060;

### Funding Statement

This study was funded by Novavax, Inc. Samples from 2019nCoV-301 (ClinicalTrials.gov: NCT04611802) were included in the present study. 2019nCoV-301 was funded by the Office of the Assistant Secretary for Preparedness and Response, Biomedical Advanced Research and Development Authority (BARDA; contract Operation Warp Speed: Novavax Project Agreement number 1 under Medical CBRN [Chemical, Biological, Radiological, and Nuclear] Defense Consortium base agreement no. 2020-530; Department of Defense no. W911QY20C0077); and the National Institute of Allergy and Infectious Diseases (NIAID), National Institutes of Health. The NIAID provides grant funding to the HIV Vaccine Trials Network (HVTN) Leadership and Operations Center (UM1 AI68614), the HVTN Statistics and Data Management Center (UM1 AI68635), the HVTN Laboratory Center (UM1 AI68618), the HIV Prevention Trials Network Leadership and Operations Center (UM1 AI68619), the AIDS Clinical Trials Group Leadership and Operations Center (UM1 AI68636), and the Infectious Diseases Clinical Research Consortium leadership group (UM1AI148684). Samples from 2019nCoV-307 (ClinicalTrials.gov: NCT05463068) were included in the present study. 2019nCoV-307 was funded by Novavax, Inc. with support from the US BARDA (Contract W15QKN-16-9-1002, Project Number MCDC2011-001).

### Summary of Updates

Dear medRxiv editorial office, In this revision, we have revised Figure 3 and text in Figure 4 legend (MN50 to ID50) in the main manuscript. We also revised the supplementary materials by replacing "trimer" to "RBD" in the legends for Figures s4c, s4d, and s4e, and replaced figure s3c for the same reasons as Figure 3 in the main manuscript. Thank you. Best, Raj Kalkeri

## References

1. Pillai S. Is it bad, is it good, or is IgG4 just misunderstood? Sci Immunol. Mar 31 2023;8(81):eadg7327. doi:10.1126/sciimmunol.adg7327

2. Rispens T, Huijbers MG. The unique properties of IgG4 and its roles in health and disease. Nat Rev Immunol. Nov 2023;23(11):763–778. doi:10.1038/s41577-023-00871-z

3. Vidarsson G, Dekkers G, Rispens T. IgG subclasses and allotypes: from structure to effector functions. Front Immunol. 2014;5:520. doi:10.3389/fimmu.2014.00520

4. Akhtar M, Islam MR, Khaton F, et al. Appearance of tolerance-induction and non-inflammatory SARS-CoV-2 spike-specific IgG4 antibodies after COVID-19 booster vaccinations. Front Immunol. 2023;14:1309997. doi:10.3389/fimmu.2023.1309997

5. Buhre JS, Pongracz T, Kunsting I, et al. mRNA vaccines against SARS-CoV-2 induce comparably low long-term IgG Fc galactosylation and sialylation levels but increasing long-term IgG4 responses compared to an adenovirus-based vaccine. Front Immunol. 2022;13:1020844. doi:10.3389/fimmu.2022.1020844

6. Espino AM, Armina-Rodriguez A, Alvarez L, et al. The Anti-SARS-CoV-2 IgG1 and IgG3 Antibody Isotypes with Limited Neutralizing Capacity against Omicron Elicited in a Latin Population a Switch toward IgG4 after Multiple Doses with the mRNA Pfizer-BioNTech Vaccine. Viruses. Jan 26 2024;16(2)doi:10.3390/v16020187

7. Hartley GE, Fryer HA, Gill PA, et al. Homologous but not heterologous COVID-19 vaccine booster elicits IgG4+ B-cells and enhanced Omicron subvariant binding. NPJ Vaccines. Jul 17 2024;9(1):129. doi:10.1038/s41541-024-00919-8

8. Irrgang P, Gerling J, Kocher K, et al. Class switch toward noninflammatory, spike-specific IgG4 antibodies after repeated SARS-CoV-2 mRNA vaccination. Sci Immunol. Jan 27 2023;8(79):eade2798. doi:10.1126/sciimmunol.ade2798

9. Jain S, Kumar S, Lai L, et al. XBB.1.5 monovalent booster improves antibody binding and neutralization against emerging SARS-CoV-2 Omicron variants. bioRxiv. Feb 5 2024;doi:10.1101/2024.02.03.578771

10. Kalkeri R. Distinct Differences in IgG4 Switch and Fc Effector Functions Between mRNA and Novavax Protein-based SARS-CoV-2 Vaccines. presented at: World Vaccine Congress; 2024; Washington, DC.

11. Kiszel P, Sik P, Miklos J, et al. Class switch towards spike protein-specific IgG4 antibodies after SARS-CoV-2 mRNA vaccination depends on prior infection history. Sci Rep. Aug 13 2023;13(1):13166. doi:10.1038/s41598-023-40103-x

12. Kobbe R. Delayed induction of non-inflammatory SARS-CoV-2 spike-specific IgG4 antibodies detected one year after BNT162b2 vaccination in children. 2024:

13. Lasrado N, Collier AY, Miller J, et al. Waning immunity and IgG4 responses following bivalent mRNA boosting. Sci Adv. Feb 23 2024;10(8):eadj9945. doi:10.1126/sciadv.adj9945

14. Reinig S, Kuo C, Wu CC, Huang SY, Yu JS, Shih SR. Specific long-term changes in anti-SARS-CoV-2 IgG modifications and antibody functions in mRNA, adenovector, and protein subunit vaccines. J Med Virol. Jul 2024;96(7):e29793. doi:10.1002/jmv.29793

15. Routhu NK, Stampfer SD, Lai L, et al. Efficacy of mRNA-1273 and Novavax ancestral or BA.1 spike booster vaccines against SARS-CoV-2 BA.5 infection in nonhuman primates. Sci Immunol. Oct 27 2023;8(88):eadg7015. doi:10.1126/sciimmunol.adg7015

16. Selva KJ, Ramanathan P, Haycroft ER, et al. Preexisting immunity restricts mucosal antibody recognition of SARS-CoV-2 and Fc profiles during breakthrough infections. JCI Insight. Sep 22 2023;8(18)doi:10.1172/jci.insight.172470

17. Sheehan J, Ardizzone CM, Khanna M, Trauth AJ, Hagensee ME, Ramsay AJ. Dynamics of Serum-Neutralizing Antibody Responses in Vaccinees through Multiple Doses of the BNT162b2 Vaccine. Vaccines (Basel). Nov 15 2023;11(11)doi:10.3390/vaccines11111720

18. Yoshimura M, Sakamoto A, Ozuru R, et al. The appearance of anti-spike receptor binding domain immunoglobulin G4 responses after repetitive immunization with messenger RNA-based COVID-19 vaccines. Int J Infect Dis. Nov 27 2023;139:1–5. doi:10.1016/j.ijid.2023.11.028

19. Valk AM, Keijser JBD, van Dam KPJ, et al. Suppressed IgG4 class switching in dupilumab- and TNF inhibitor-treated patients after mRNA vaccination. Allergy. Jul 2024;79(7):1952–1961. doi:10.1111/all.16089

20. Marchese AM, Fries L, Beyhaghi H, et al. Mechanisms and implications of IgG4 responses to SARS-CoV-2 and other repeatedly administered vaccines. J Infect. Oct 16 2024;89(6):106317. doi:10.1016/j.jinf.2024.106317

21. Bruhns P, Iannascoli B, England P, et al. Specificity and affinity of human Fcgamma receptors and their polymorphic variants for human IgG subclasses. Blood. Apr 16 2009;113(16):3716–25. doi:10.1182/blood-2008-09-179754

22. Smith KG, Clatworthy MR. FcgammaRIIB in autoimmunity and infection: evolutionary and therapeutic implications. Nat Rev Immunol. May 2010;10(5):328–43. doi:10.1038/nri2762

23. Willcocks LC, Smith KG, Clatworthy MR. Low-affinity Fcgamma receptors, autoimmunity and infection. Expert Rev Mol Med. Aug 13 2009;11:e24. doi:10.1017/S1462399409001161

24. Reinig S, Shih SR. Non-neutralizing functions in anti-SARS-CoV-2 IgG antibodies. Biomed J. Feb 2024;47(1):100666. doi:10.1016/j.bj.2023.100666

25. Lu LL, Suscovich TJ, Fortune SM, Alter G. Beyond binding: antibody effector functions in infectious diseases. Nat Rev Immunol. Jan 2018;18(1):46–61. doi:10.1038/nri.2017.106

26. Zhang W, Chua BY, Selva KJ, et al. SARS-CoV-2 infection results in immune responses in the respiratory tract and peripheral blood that suggest mechanisms of disease severity. Nat Commun. May 19 2022;13(1):2774. doi:10.1038/s41467-022-30088-y

27. Kalkeri R, Zhu M, Cloney-Clark S, et al. Altered IgG4 antibody response to repeated mRNA versus recombinant protein SARS-CoV-2 vaccines. J Infect. Feb 13 2024;88(3):106119. doi:10.1016/j.jinf.2024.106119

28. Kober C, Manni S, Wolff S, et al. IgG3 and IgM Identified as Key to SARS-CoV-2 Neutralization in Convalescent Plasma Pools. PLoS One. 2022;17(1):e0262162. doi:10.1371/journal.pone.0262162

29. Goldblatt D, Alter G, Crotty S, Plotkin SA. Correlates of protection against SARS-CoV-2 infection and COVID-19 disease. Immunol Rev. Sep 2022;310(1):6–26. doi:10.1111/imr.13091

30. Bartsch YC, Tong X, Kang J, et al. Omicron variant Spike-specific antibody binding and Fc activity are preserved in recipients of mRNA or inactivated COVID-19 vaccines. Sci Transl Med. Apr 27 2022;14(642):eabn9243. doi:10.1126/scitranslmed.abn9243

31. Kaplonek P, Fischinger S, Cizmeci D, et al. mRNA-1273 vaccine-induced antibodies maintain Fc effector functions across SARS-CoV-2 variants of concern. Immunity. Feb 8 2022;55(2):355-365 e4. doi:10.1016/j.immuni.2022.01.001

32. Richardson SI, Madzorera VS, Spencer H, et al. SARS-CoV-2 Omicron triggers cross-reactive neutralization and Fc effector functions in previously vaccinated, but not unvaccinated, individuals. Cell Host Microbe. Jun 8 2022;30(6):880–886 e4. doi:10.1016/j.chom.2022.03.029

33. Fong Y, Huang Y, Benkeser D, et al. Immune correlates analysis of the PREVENT-19 COVID-19 vaccine efficacy clinical trial. Nat Commun. Jan 19 2023;14(1):331. doi:10.1038/s41467-022-35768-3

34. Fong Y, McDermott AB, Benkeser D, et al. Immune correlates analysis of the ENSEMBLE single Ad26.COV2.S dose vaccine efficacy clinical trial. Nat Microbiol. Dec 2022;7(12):1996-2010. doi:10.1038/s41564-022-01262-1

35. Gilbert PB, Montefiori DC, McDermott AB, et al. Immune correlates analysis of the mRNA-1273 COVID-19 vaccine efficacy clinical trial. Science. Jan 7 2022;375(6576):43-50. doi:10.1126/science.abm3425

36. Tortorici MA, Addetia A, Seo AJ, et al. Persistent immune imprinting occurs after vaccination with the COVID-19 XBB.1.5 mRNA booster in humans. Immunity. Apr 9 2024;57(4):904-911.e4. doi:10.1016/j.immuni.2024.02.016

37. Dunkle LM, Kotloff KL, Gay CL, et al. Efficacy and Safety of NVX-CoV2373 in Adults in the United States and Mexico. N Engl J Med. Feb 10 2022;386(6):531-543. doi:10.1056/NEJMoa2116185

38. Raiser F, Davis M, Adelglass J, et al. Immunogenicity and safety of NVX-CoV2373 as a booster: A phase 3 randomized clinical trial in adults. Vaccine. Sep 22 2023;41(41):5965-5973. doi:10.1016/j.vaccine.2023.07.056

39. Babu TM, Shen X, McClelland RS, et al. Severe Acute Respiratory Syndrome Coronavirus 2 Omicron Subvariant Neutralization Following a Primary Vaccine Series of NVX-CoV2373 and BNT162b2 Monovalent Booster Vaccine. Open Forum Infect Dis. Feb 2024;11(2):ofad673. doi:10.1093/ofid/ofad673

40. Zhu M, Cloney-Clark S, Feng SL, et al. A Severe Acute Respiratory Syndrome Coronavirus 2 Anti-Spike Immunoglobulin G Assay: A Robust Method for Evaluation of Vaccine Immunogenicity Using an Established Correlate of Protection. Microorganisms. Jul 11 2023;11(7)doi:10.3390/microorganisms11071789

41. Hamilton S, Zhu M, Cloney-Clark S, et al. Validation of a severe acute respiratory syndrome coronavirus 2 microneutralization assay for evaluation of vaccine immunogenicity. J Virol Methods. Jun 2024;327:114945. doi:10.1016/j.jviromet.2024.114945

42. Selva KJ, van de Sandt CE, Lemke MM, et al. Systems serology detects functionally distinct coronavirus antibody features in children and elderly. Nat Commun. Apr 1 2021;12(1):2037. doi:10.1038/s41467-021-22236-7

43. Lopez E, Haycroft ER, Adair A, et al. Simultaneous evaluation of antibodies that inhibit SARS-CoV-2 variants via multiplex assay. JCI Insight. Aug 23 2021;6(16)doi:10.1172/jci.insight.150012

44. Brown EP, Licht AF, Dugast AS, et al. High-throughput, multiplexed IgG subclassing of antigen-specific antibodies from clinical samples. J Immunol Methods. Dec 14 2012;386(1-2):117–23. doi:10.1016/j.jim.2012.09.007

45. Cai Z, Kalkeri R, Wang M, et al. Validation of a Pseudovirus Neutralization Assay for Severe Acute Respiratory Syndrome Coronavirus 2: A High-Throughput Method for the Evaluation of Vaccine Immunogenicity. Microorganisms. Jun 14 2024; 12(6):1201. doi: 10.3390/microorganisms12061201.

