## Supplementary File for "Anti-Spike IgG4 and Fc Effector Responses: The Impact of SARS-CoV-2 Vaccine Platform–Specific Priming and Immune Imprinting"

**Running title**: COVID-19 Vaccine Priming and IgG4

**Corresponding author:**

Raj Kalkeri, PhD

Novavax, Inc., Gaithersburg, MD, USA

**Supplemental Figures:**

**Fig.s1a:** Anti-Spike (rS) IgG isotypes in **anti-nucleoprotein reactive (Anti-NP positive)** subjects primed either with mRNA vaccines (mRNA-1273 or BNT162b2) or the protein-based subunit COVID-19 vaccine (NVX-CoV2373). Pre-booster (day 0), post-booster NVX-CoV2373 (day 28) or mRNA (BNT162b2) vaccines (day 29). ELISA based quantitation [Geometric Mean Titers (GMT) with 95% confidence intervals (CI)] of IgG isotypes is shown in **A–D**. **A–D:** Gray – mRNA vaccines x3 +NVX-CoV2373 (N=8), dark-blue – NVX-CoV273 x3 + NVX-CoV273 (N=8), **E–H:** light-blue – NVX-CoV273 x2 + BNT162b2 (N=5 for Day 0 and N=7 for Day 29, except for IgG4 N=6 for Day 29). Statistical analysis was performed by the two-tailed Mann–Whittney test. *p<0.05, **p < 0.01 ***p < 0.001 indicate significance. n.s. indicates not significant.

**Fig.s1b:** Anti-Spike (rS) IgG isotypes in **anti-nucleoprotein non-reactive (Anti-NP negative)** subjects primed either with mRNA vaccines (mRNA-1273 or BNT162b2) or the protein-based subunit COVID-19 vaccine (NVX-CoV2373). Pre-booster (day 0), post-booster NVX-CoV2373 (day 28) or mRNA (BNT162b2) vaccines (day 29). ELISA based quantitation [Geometric Mean Titers (GMT) with 95% confidence intervals (CI)] of IgG isotypes is shown in **A–D**. **A–D:** Gray – mRNA vaccines x3 +NVX-CoV2373 (N=10), dark-blue – NVX-CoV273 x3 + NVX-CoV273 (N=10), **E–H:** light-blue – NVX-CoV273 x2 + BNT162b2 (N=19 for Day 0 and N=23 for Day 29). Statistical analysis was performed by the two-tailed Mann–Whittney test. * p<0.05, **p < 0.01 ***p < 0.001, ****p<0.0001 indicate significance. n.s. indicates not significant.

**Fig.s2a:** Fc effector assay (surrogate assays for ADCP, ADCC, and ADCD) data in **anti-nucleoprotein reactive (Anti-NP–positive)** subjects primed either with mRNA vaccines (mRNA-1273 or BNT162b2) or the protein-based subunit COVID-19 vaccine (NVX-CoV2373). Pre-booster (day 0), post-booster NVX-CoV2373 (day 28) or mRNA (BNT162b2) vaccines (day 29). GMT with 95% CI is shown. **A, D.** Surrogate assays for ADCP (FcgR2a binding), **B, E.** Surrogate assays for ADCC (FcgR3a binding), **C, F.** Surrogate assays for ADCD (C1q binding). **A–C:** Gray – mRNA vaccines x3 +NVX-CoV2373 (N=8), dark-blue – NVX-CoV273 x3 + NVX-CoV273 (N=7), **D–F:** light-blue – NVX-CoV273 x2 + BNT162b2 (N=5 for Day 0 and N=7 for Day 29). Statistical analysis was performed by the two-tailed Mann–Whittney test. *p < 0.05, **p < 0.01 indicate significance. n.s. indicates not significant.

**Fig.s2b:** Fc effector assay (surrogate assays for ADCP, ADCC, ADCD) data in **anti-nucleoprotein non-reactive (anti-NP–negative)** subjects primed either with mRNA vaccines (mRNA-1273 or BNT162b2) or the protein-based subunit COVID-19 vaccine (NVX-CoV2373). Pre-booster (day 0), post-booster NVX-CoV2373 (day 28) or mRNA (BNT162b2) vaccines (day 29). GMT with 95% CI is shown. **A, D:** Surrogate assays for ADCP (FcgR2a binding), **B, E:** Surrogate assays for ADCC (FcgR3a binding), **C, F:** Surrogate assays for ADCD (C1q binding). **A–C:** Gray – mRNA vaccines x3 +NVX-CoV2373 (N=10), dark blue- NVX-CoV273 x3 + NVX-CoV273 (N=10), **D–F:** light blue- NVX-CoV273 x2 + BNT162b2 (N=19 for Day 0 and N=23 for Day 29). Statistical analysis was performed by the two-tailed Mann–Whittney test. *p < 0.05, ****p < 0.0001, indicate significance. n.s. indicates “not significant”.

**Fig.s3c:** Anti-Spike (rS) IgG isotype proportions in **all subjects [(anti-nucleoprotein non-reactive (anti-NP–negative) and anti-nucleoprotein reactive (anti-NP–positive)]** receiving the protein-based subunit XBB.1.5 COVID-19 vaccine (NVX-CoV2601) after priming with mRNA vaccines (≥3x). Pre-booster (day 0), post-booster with the NVX-CoV2601 vaccine (day 28). **A.** Anti-rS IgG isotype proportions (pie chart) against the wild-type (WT) **RBD antigen** (ancestral) **B.** Anti-rS IgG isotype proportions (pie chart) against the XBB.1.5 **RBD antigen C.** Anti-rS IgG isotype levels (GMT with 95% CI bar graph) against WT (Stripes, N=29) and XBB.1.5 (Solid color, N=29) **RBD antigens**. Geometric mean fold raise (GMFR) at day 28 post-booster compared to pre-booster levels (day 0) are shown above the bars. Light-gray – anti-rS IgG1, medium-gray – anti-rS IgG2, black – anti-rS IgG3, plum – anti-rS IgG4. Statistical analysis was performed by the two-tailed Mann–Whittney test. ****p < 0.0001.

**Fig.s3d:** Anti-Spike (rS) IgG isotype proportions in **seropositive subjects [anti-nucleoprotein reactive (anti-NP–positive)]** receiving the protein-based subunit XBB.1.5 COVID-19 vaccine (NVX-CoV2601) after priming with mRNA vaccines (≥3x). Pre-booster (day 0), post-booster with the NVX-CoV2601 vaccine (day 28). **A.** Anti-rS IgG isotype proportions (pie chart) against the wild-type (WT) **RBD antigen** (ancestral) **B.** Anti-rS IgG isotype proportions (pie chart) against the XBB.1.5 **RBD antigen C.** Anti-rS IgG isotype levels (GMT with 95% CI bar graph) against WT (Stripes, N=19) and XBB.1.5 (Solid color, N=19) **RBD antigens**. Geometric mean fold raise (GMFR) at day 28 post-booster compared to pre-booster levels (day 0) are shown above the bars. Light-gray – anti-rS IgG1, medium-gray – anti-rS IgG2, black – anti-rS IgG3, plum – anti-rS IgG4. Statistical analysis was performed by the two-tailed Mann–Whittney test. *** p<0.001 and ****p < 0.0001.

**Fig.s3e:** Anti-Spike (rS) IgG isotype proportions in **seronegative subjects [anti-nucleoprotein non-reactive (anti-NP–negative)]** receiving the protein-based subunit XBB.1.5 COVID-19 vaccine

(NVX-CoV2601) after priming with mRNA vaccines (≥3x). Pre-booster (day 0), post-booster with the NVX-CoV2601 vaccine (day 28). **A.** Anti-rS IgG isotype proportions (pie chart) against the wild-type (WT) **RBD antigen** (ancestral) **B.** Anti-rS IgG isotype proportions (pie chart) against the XBB.1.5 **RBD antigen C.** Anti-rS IgG isotype levels (GMT with 95% CI bar graph) against WT (Stripes, N=10) and XBB.1.5 (Solid color, N=10) **RBD antigens**. Geometric mean fold raise (GMFR) at day 28 post-booster compared to pre-booster levels (day 0) are shown above the bars. Light-gray – anti-rS IgG1, medium-gray – anti-rS IgG2, black – anti-rS IgG3, plum – anti-rS IgG4. Statistical analysis was performed by the two-tailed Mann–Whittney test. ***p<0.001, ****p < 0.0001.

**Fig.s4a:** Neutralizing antibody levels and Fc effector assay data (surrogate assays for ADCP, ADCC, and ADCD) in **anti-nucleoprotein reactive (anti-NP–positive)** subjects receiving the protein-based subunit XBB.1.5 COVID-19 vaccine (NVX-CoV2601) after priming with mRNA vaccines (≥3x). Pre-booster (day 0), post-booster with the NVX-CoV2601 vaccine (day 28). **A**. Neutralizing antibody titers against ancestral (WT) (N=19) and XBB.1.5 (N=19) strains measured by pseudovirus neutralization assay **B.** Surrogate assays for ADCP (FcgR2a binding), against ancestral (WT) (N=19) and XBB.1.5 (N=19) trimer antigens **C.** Surrogate assays for ADCC (FcgR3a binding), against ancestral (WT) (N=19) and XBB.1.5 (N=19) trimer antigens. **D.** C1q binding for ADCD, against ancestral (WT) (N=19) and XBB.1.5 (N=19) trimer antigens. GMT with 95% CI is shown. Geometric mean fold raise (GMFR) at day 28 post-booster compared to pre-booster levels (day 0) are shown above the bars. Gray stripes – ancestral, red – XBB.1.5. Statistical analysis was performed by the two-tailed Mann–Whittney test. **p<0.01, ***p<0.001. n.s. indicates "not significant".

**Fig.s4b:** Neutralizing antibody levels and Fc effector assay data (surrogate assays for ADCP, ADCC, and ADCD) in **anti-nucleoprotein non-reactive (anti-NP–negative)** subjects receiving the protein-based subunit XBB.1.5 COVID-19 vaccine (NVX-CoV2601) after priming with mRNA vaccines (≥3x). Pre-booster (day 0), post-booster with the NVX-CoV2601 vaccine (day 28). **A**. Neutralizing antibody titers against ancestral (WT) (N=10) and XBB.1.5 (N=10) strains measured by pseudovirus neutralization assay **B.** Surrogate assays for ADCP (FcgR2a binding), against ancestral (WT) (N=10) and XBB.1.5 (N=10) trimer antigens **C.** Surrogate assays for ADCC (FcgR3a binding), against ancestral (WT) (N=10) and XBB.1.5 (N=10) trimer antigens. **D.** C1q binding for ADCD, against ancestral (WT) (N=10) and XBB.1.5 (N=10) trimer antigens. GMT with 95% CI is shown. Geometric mean fold raise (GMFR) at day 28 post-booster compared to pre-booster levels (day 0) are shown above the bars. Gray stripes – ancestral, red – XBB.1.5. Statistical analysis was performed by the two-tailed Mann–Whittney test. *p < 0.05. **p<0.01, n.s. indicates "not significant".

**Fig.s4c:** Fc effector assay data (surrogate assays for ADCP, ADCC, and ADCD) in **all subjects [(anti-nucleoprotein non-reactive (anti-NP–negative) and anti-nucleoprotein reactive (anti-NP–positive)]** subjects receiving the protein-based subunit XBB.1.5 COVID-19 vaccine (NVX-CoV2601) after priming with mRNA vaccines (≥3x). Pre-booster (day 0), post-booster with the NVX-CoV2601 vaccine (day 28). **A.** Surrogate assays for ADCP (FcgR2a binding), against ancestral (WT) (N=29) and XBB.1.5 (N=29) **RBD** antigens **B.** Surrogate assays for ADCC (FcgR3a binding), against ancestral (WT) (N=29) and XBB.1.5 (N=29) **RBD** antigens. **C.** C1q binding for ADCD, against WT (N=29) and XBB.1.5 (N=29) **RBD** antigens. GMT with 95% CI is shown. Geometric mean fold raise (GMFR) at day 28 post-booster compared to pre-booster levels (day 0) are shown above the bars. Gray stripes – ancestral, red – XBB.1.5. Statistical analysis was performed by the two-tailed Mann–Whittney test. ****p < 0.0001.

**Fig.s4d:** Fc effector assay data (surrogate assays for ADCP, ADCC, and ADCD) in **seropositive subjects [anti-nucleoprotein reactive (anti-NP–positive)]** subjects receiving the protein-based subunit XBB.1.5 COVID-19 vaccine (NVX-CoV2601) after priming with mRNA vaccines (≥3x). Pre-booster (day 0), post-booster with the NVX-CoV2601 vaccine (day 28). **A.** Surrogate assays for ADCP (FcgR2a binding), against ancestral (WT) (N=19) and XBB.1.5 (N=19) **RBD** antigens **B.** Surrogate assays for ADCC (FcgR3a binding), against ancestral (WT) (N=19) and XBB.1.5 (N=19) **RBD** antigens. **C.** C1q binding for ADCD, against WT (N=19) and XBB.1.5 (N=19) **RBD** antigens. GMT with 95% CI is shown. Geometric mean fold raise (GMFR) at day 28 post-booster compared to pre-booster levels (day 0) are shown above the bars. Gray stripes – ancestral, red – XBB.1.5. Statistical analysis was performed by the two-tailed Mann–Whittney test. ****p < 0.0001.

**Fig.s4e:** Fc effector assay data (surrogate assays for ADCP, ADCC, and ADCD) in **seronegative subjects [(anti-nucleoprotein non-reactive (anti-NP–negative)]** subjects receiving the protein-based subunit XBB.1.5 COVID-19 vaccine (NVX-CoV2601) after priming with mRNA vaccines (≥3x). Pre-booster (day 0), post-booster with the NVX-CoV2601 vaccine (day 28). **A.** Surrogate assays for ADCP (FcgR2a binding), against ancestral (WT) (N=10) and XBB.1.5 (N=10) **RBD** antigens **B.** Surrogate assays for ADCC (FcgR3a binding), against ancestral (WT) (N=10) and XBB.1.5 (N=10) **RBD** antigens. **C.** C1q binding for ADCD, against WT (N=10) and XBB.1.5 (N=10) **RBD** antigens. GMT with 95% CI is shown. Geometric mean fold raise (GMFR) at day 28 post-booster compared to pre-booster levels (day 0) are shown above the bars. Gray stripes – ancestral, red – XBB.1.5. Statistical analysis was performed by the two-tailed Mann–Whittney test. ***p<0.001, ****p < 0.0001.

Fig. s1a

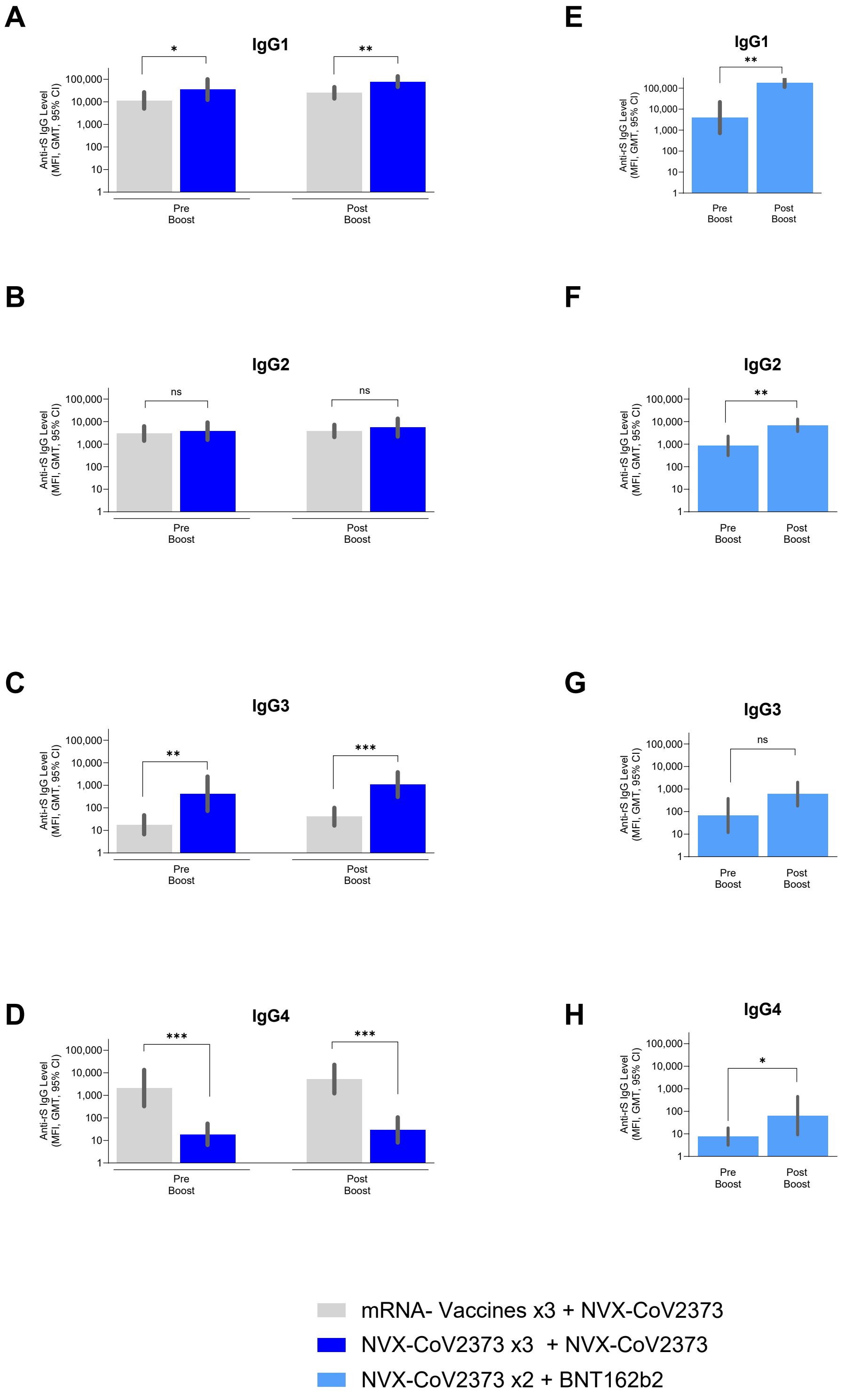
A
IgG1
*
**
Anti-rS IgG Level
(MFI, GMT, 95% CI)
100,000
10,000
1,000
100
10
1
Pre
Boost
Post
Boost
E
IgG1
**
B
IgG2
ns
ns
F
IgG2
**
C
IgG3
**
***
G
IgG3
ns
D
IgG4
***
***
H
IgG4
*
mRNA- Vaccines x3 + NVX-CoV2373
NVX-CoV2373 x3 + NVX-CoV2373
NVX-CoV2373 x2 + BNT162b2

Fig s1b

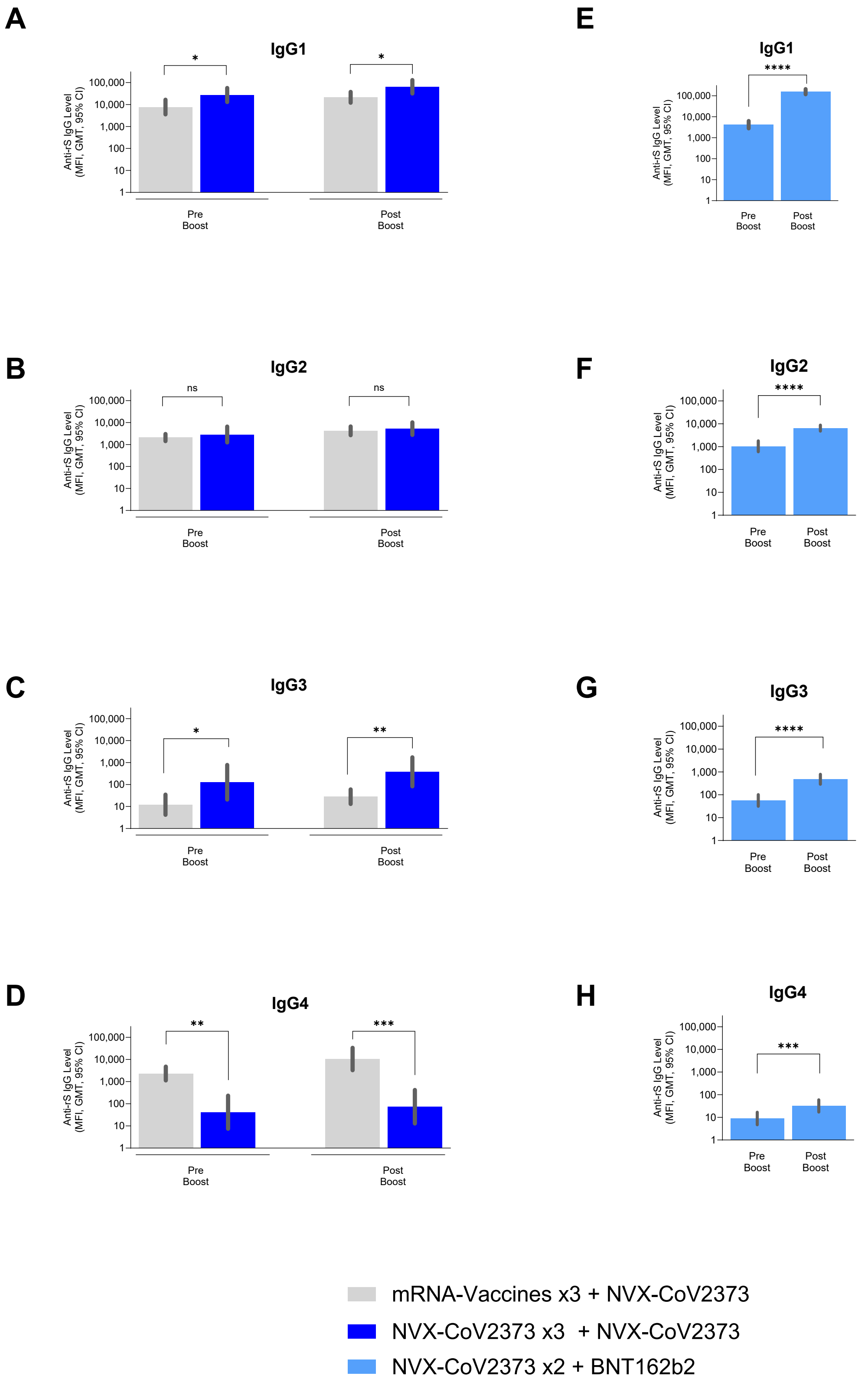

A
IgG1
*
*
Anti-rS IgG Level (MFI, GMT, 95% CI)
100,000
10,000
1,000
100
10
1
Pre Boost
Post Boost
B
IgG2
ns
ns
C
IgG3
*
**
D
IgG4
**
***
E
IgG1
****
F
IgG2
****
G
IgG3
****
H
IgG4
***
mRNA-Vaccines x3 + NVX-CoV2373
NVX-CoV2373 x3 + NVX-CoV2373
NVX-CoV2373 x2 + BNT162b2

Fig. s2a

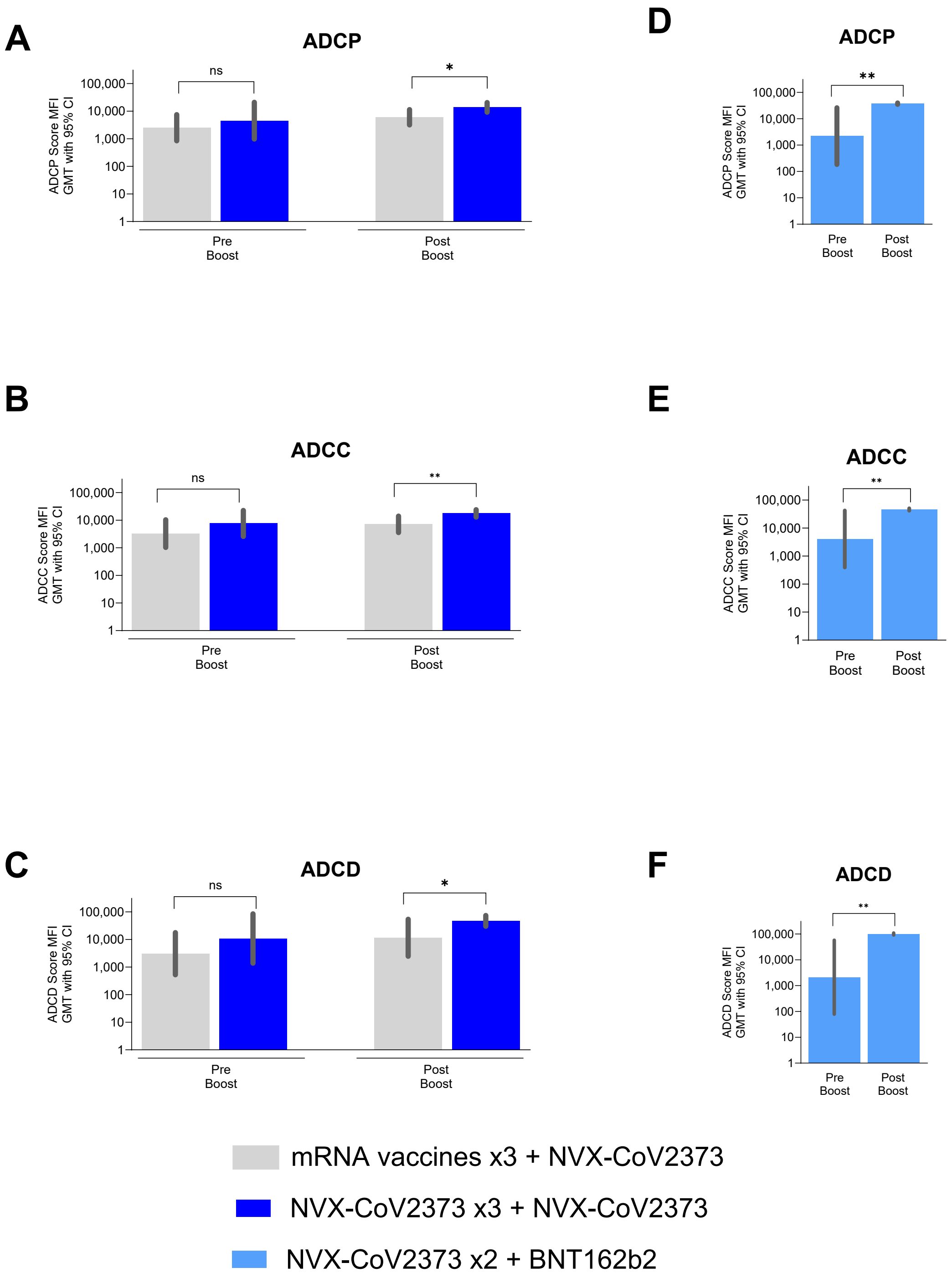

A
ADCP
ADCP Score MFI GMT with 95% CI
ns
*
Pre Boost
Post Boost
D
ADCP
**
B
ADCC
ADCC Score MFI GMT with 95% CI
ns
**
E
ADCC
C
ADCD
ADCD Score MFI GMT with 95% CI
F
ADCD
100,000
10,000
1,000
100
10
1
mRNA vaccines x3 + NVX-CoV2373
NVX-CoV2373 x3 + NVX-CoV2373
NVX-CoV2373 x2 + BNT162b2

Fig. s2b

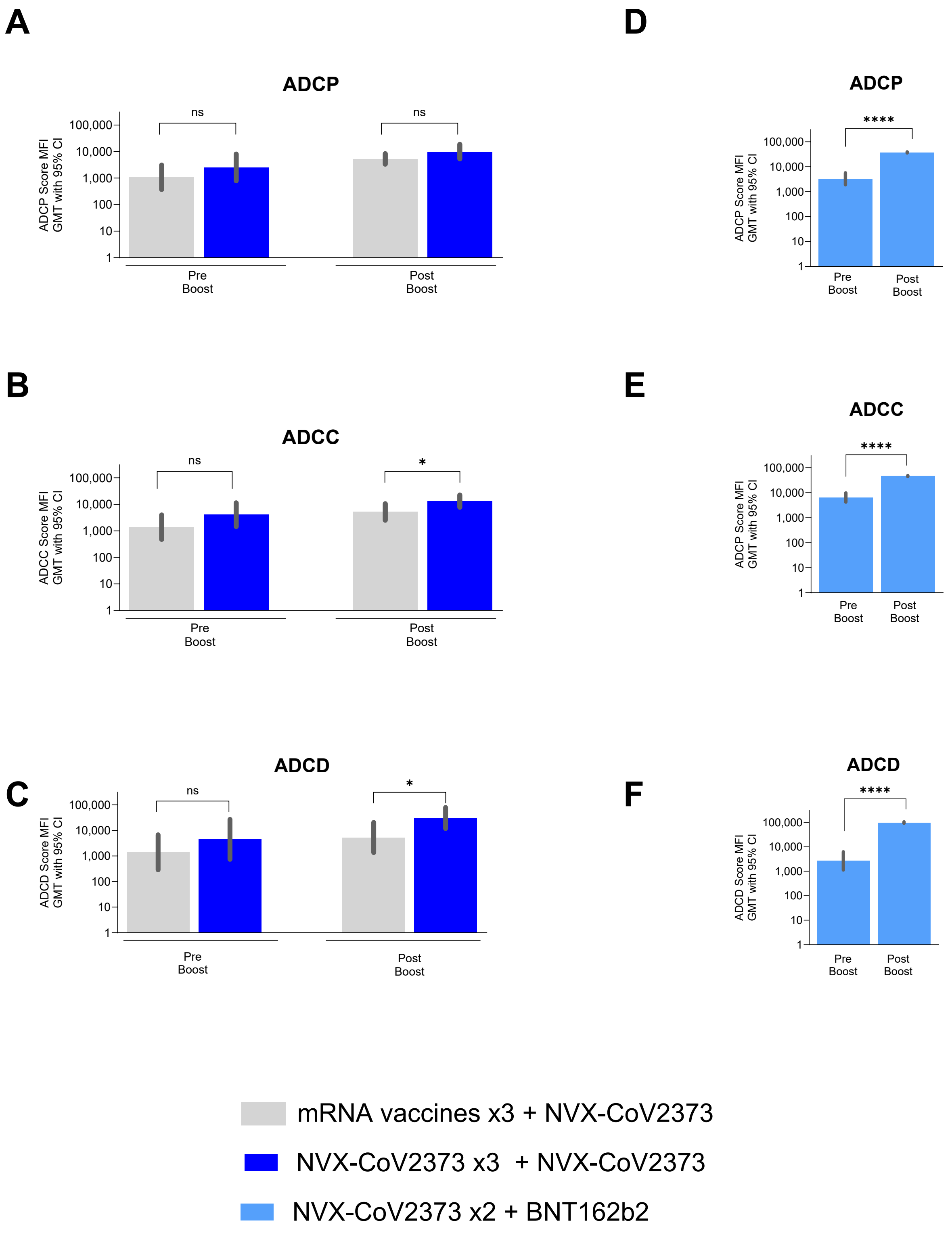
A
ADCP
ADCP Score MFI GMT with 95% CI
ns
ns
100,000
10,000
1,000
100
10
1
Pre Boost
Post Boost
B
ADCC
ADCC Score MFI GMT with 95% CI
ns
*
C
ADCD
ADCD Score MFI GMT with 95% CI
ns
*
D
ADCP
ADCP Score MFI GMT with 95% CI
****
E
ADCC
ADCP Score MFI GMT with 95% CI
****
F
ADCD
ADCD Score MFI GMT with 95% CI
****
mRNA vaccines x3 + NVX-CoV2373
NVX-CoV2373 x3 + NVX-CoV2373
NVX-CoV2373 x2 + BNT162b2

Fig.s3a

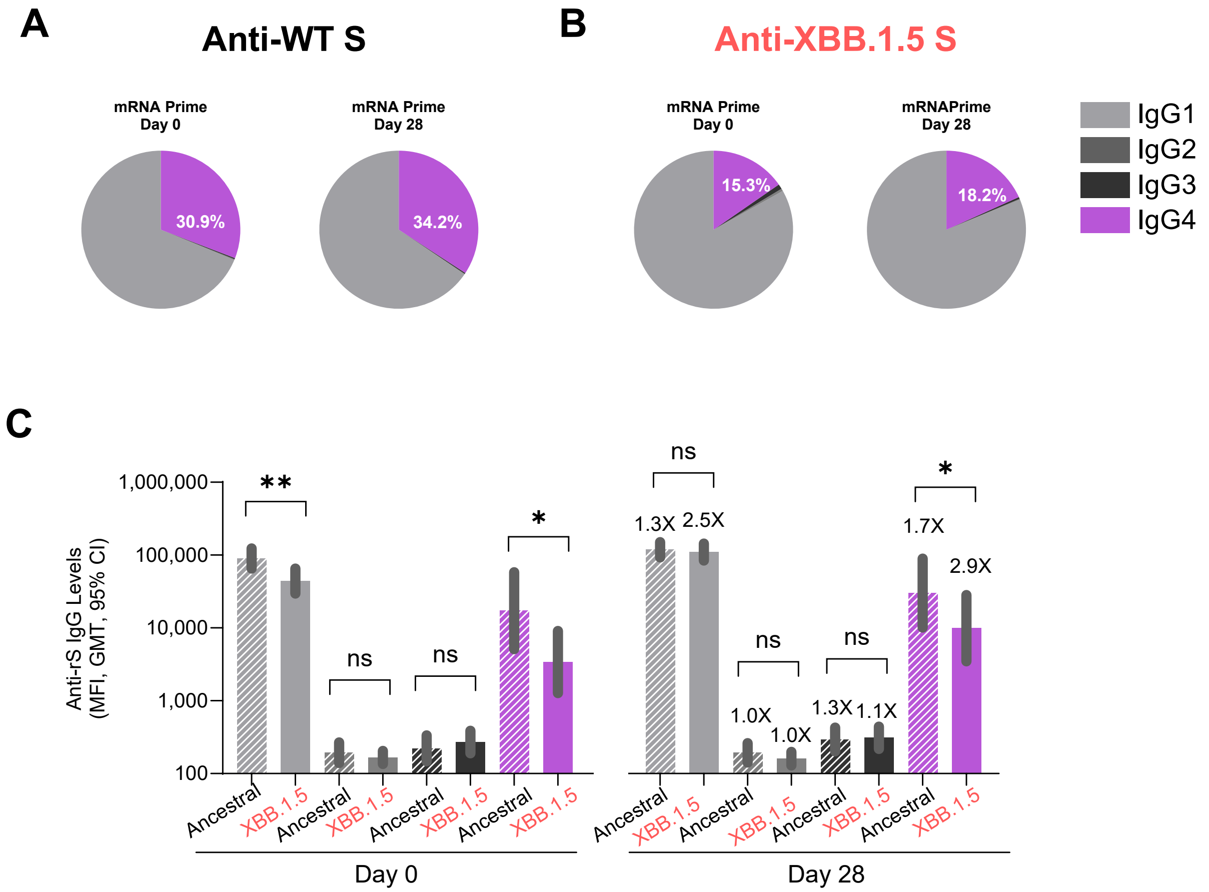
A
Anti-WT S
mRNA Prime Day 0
30.9%
mRNA Prime Day 28
34.2%
B
Anti-XBB.1.5 S
mRNA Prime Day 0
15.3%
mRNAPrime Day 28
18.2%
IgG1
IgG2
IgG3
IgG4
C
Anti-rS IgG Levels (MFI, GMT, 95% CI)
1,000,000
100,000
10,000
1,000
100
**
ns
ns
*
ns
1.3X
2.5X
ns
ns
1.0X
1.0X
1.3X
1.1X
*
1.7X
2.9X
Ancestral
XBB.1.5
Day 0
Day 28

Fig.s3b

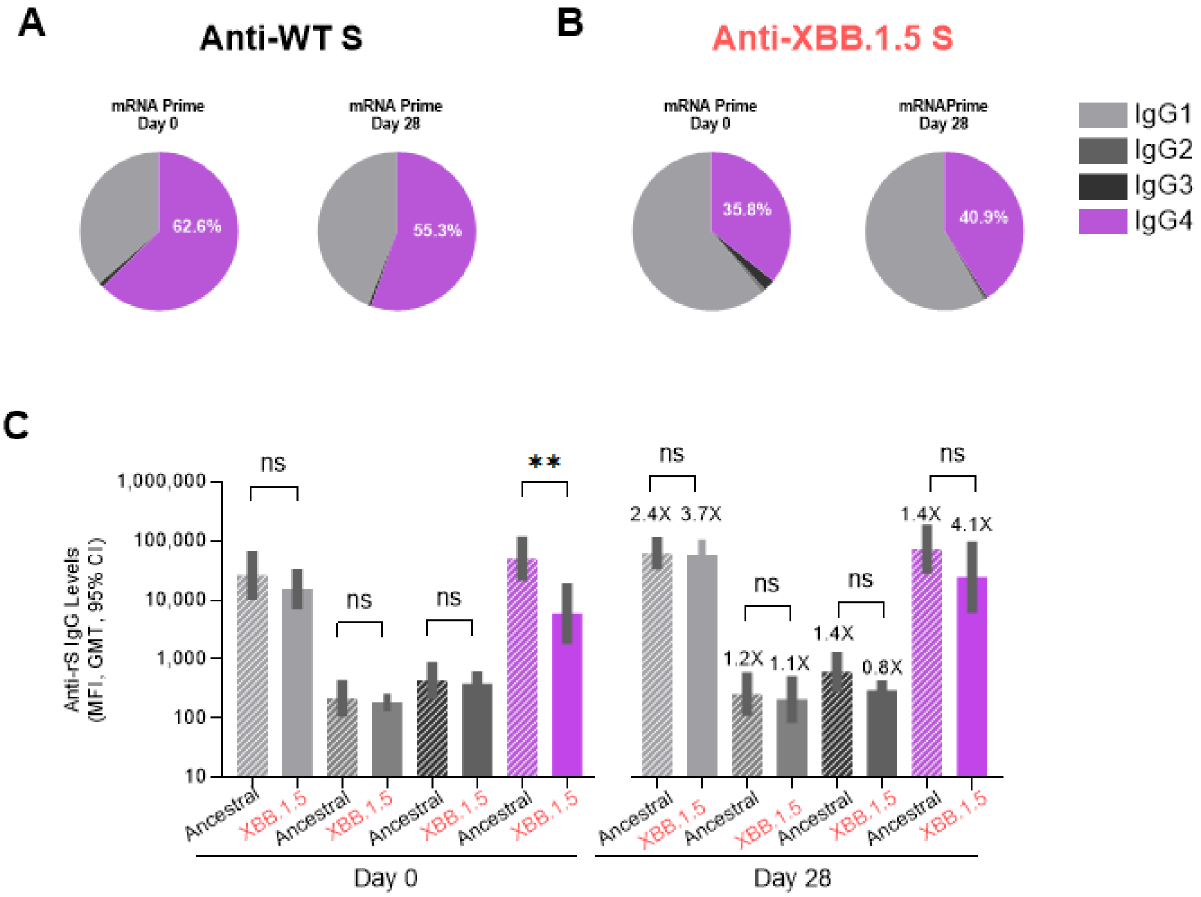

A
Anti-WT S
mRNA Prime
Day 0
62.6%
mRNA Prime
Day 28
55.3%
B
Anti-XBB.1.5 S
mRNA Prime
Day 0
35.8%
mRNAPrime
Day 28
40.9%
IgG1
IgG2
IgG3
IgG4
C
Anti-rS IgG Levels
(MFI, GMT, 95% CI)
1,000,000
100,000
10,000
1,000
100
10
ns
ns
ns
**
ns
2.4X
3.7X
ns
1.2X
1.1X
ns
1.4X
0.8X
ns
1.4X
4.1X
Ancestral
XBB.1.5
Day 0
Day 28

Fig.s3c

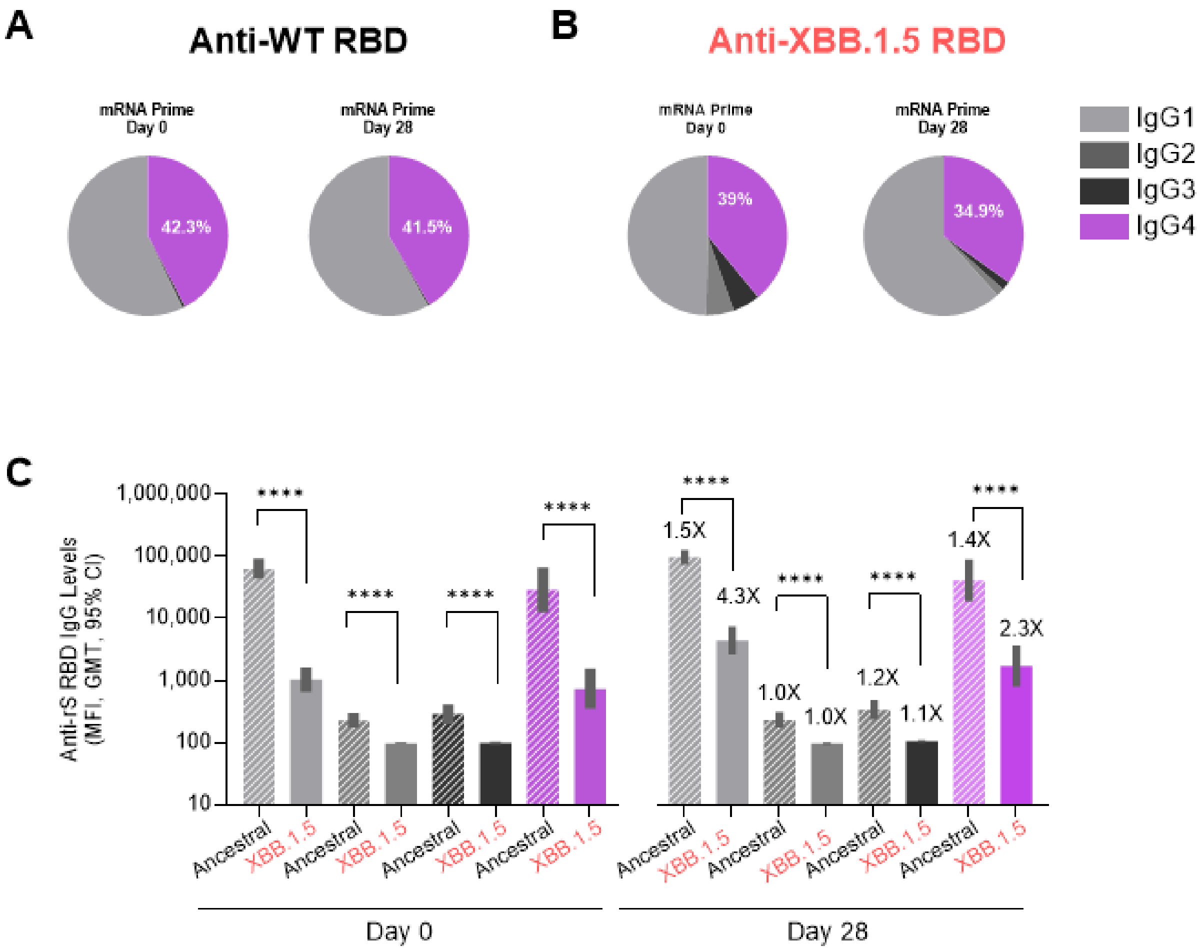
A
Anti-WT RBD
mRNA Prime
Day 0
42.3%
mRNA Prime
Day 28
41.5%
B
Anti-XBB.1.5 RBD
mRNA Prime
Day 0
39%
mRNA Prime
Day 28
34.9%
IgG1
IgG2
IgG3
IgG4
C
Anti-rS RBD IgG Levels
(MFI, GMT, 95% CI)
1,000,000
100,000
10,000
1,000
100
10
****
1.5X
4.3X
1.0X
1.0X
1.2X
1.1X
1.4X
2.3X
Ancestral
XBB.1.5
Day 0
Day 28

Fig.s3d

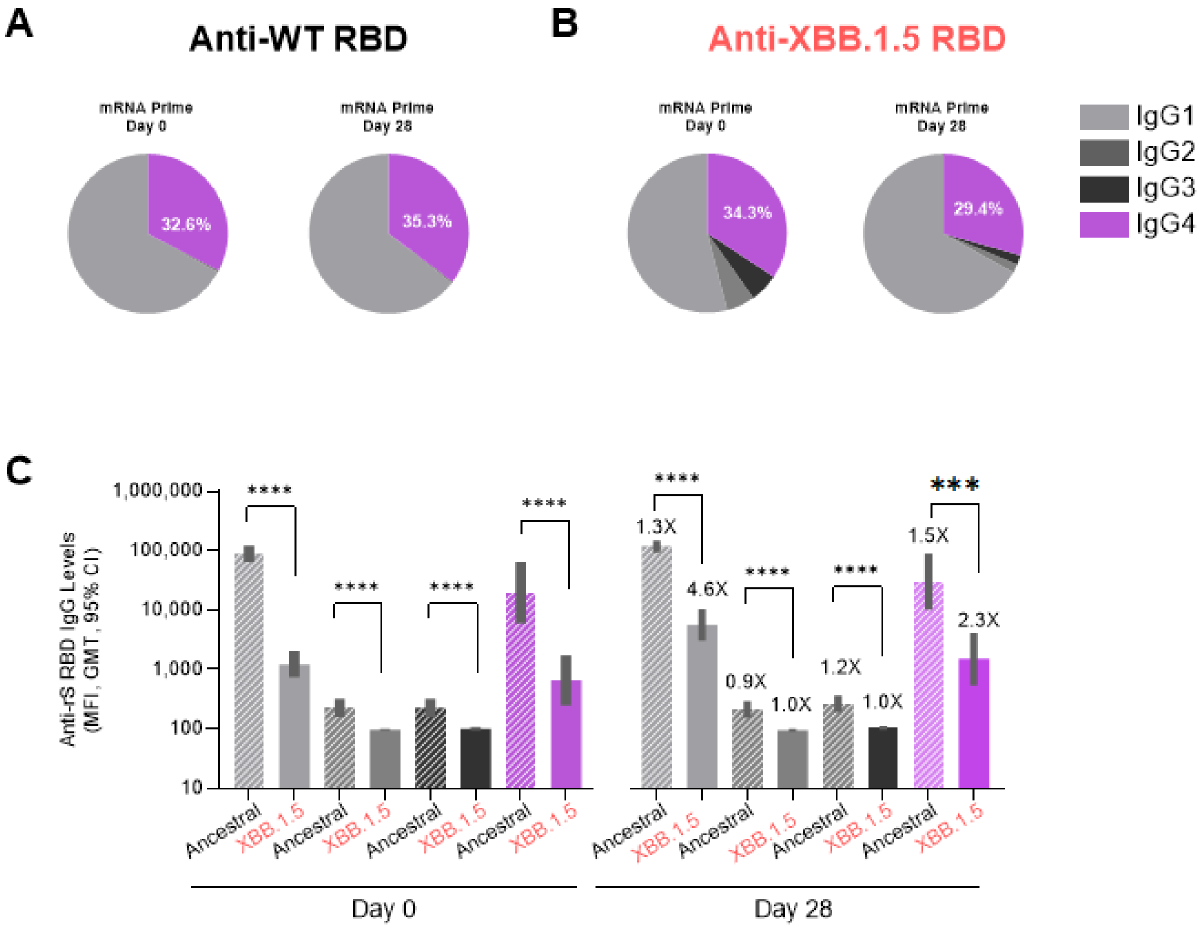
A
Anti-WT RBD
mRNA Prime
Day 0
32.6%
mRNA Prime
Day 28
35.3%
B
Anti-XBB.1.5 RBD
mRNA Prime
Day 0
34.3%
mRNA Prime
Day 28
29.4%
IgG1
IgG2
IgG3
IgG4
C
Anti-rS RBD IgG Levels
(MFI, GMT, 95% CI)
1,000,000
100,000
10,000
1,000
100
10
****
****
****
****
1.3X
4.6X
0.9X
1.0X
1.2X
1.0X
1.5X
2.3X
***
Ancestral
XBB.1.5
Day 0
Day 28

Fig.s3e

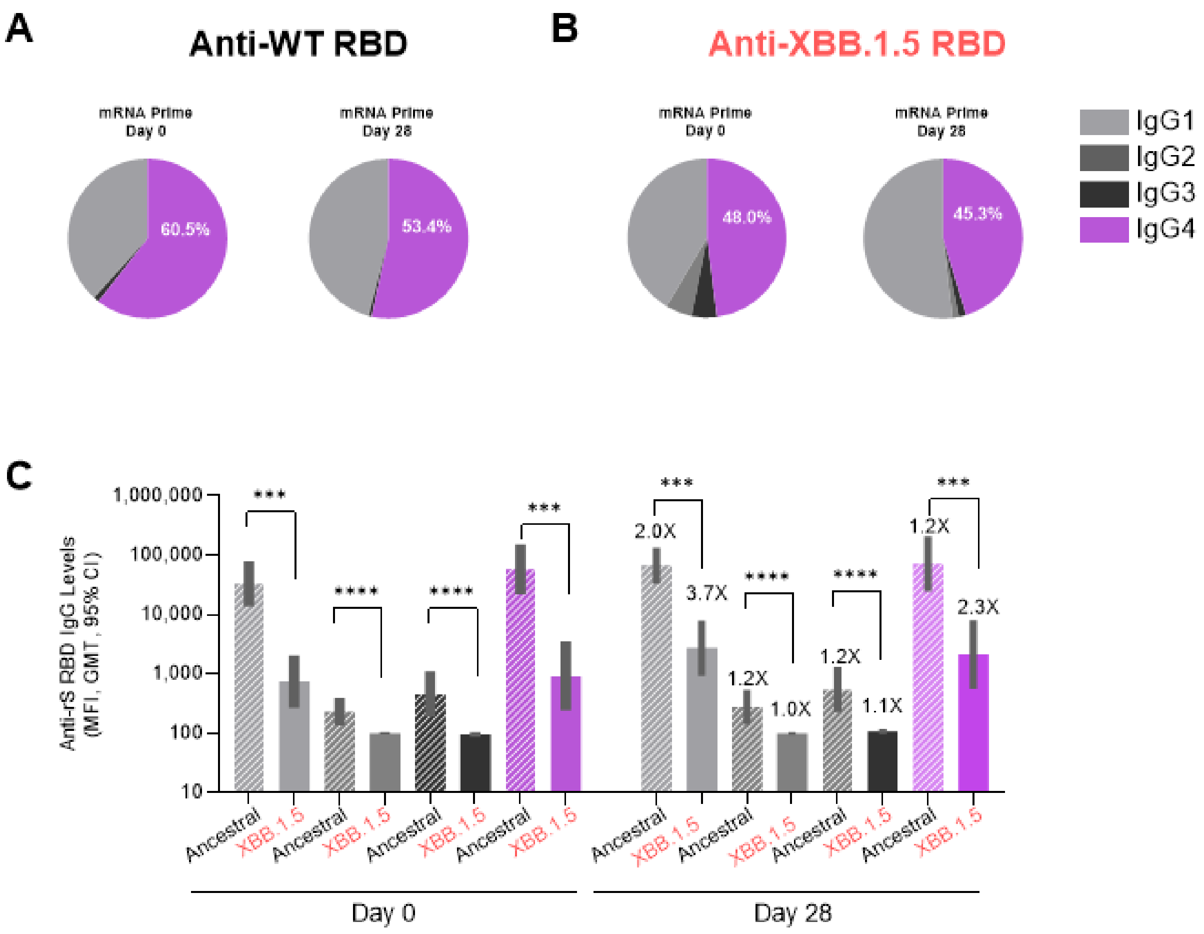
A
Anti-WT RBD
mRNA Prime Day 0
60.5%
mRNA Prime Day 28
53.4%
B
Anti-XBB.1.5 RBD
mRNA Prime Day 0
48.0%
mRNA Prime Day 28
45.3%
IgG1
IgG2
IgG3
IgG4
C
Anti-rS RBD IgG Levels (MFI, GMT, 95% CI)
1,000,000
100,000
10,000
1,000
100
10
***
****
****
***
***
****
****
***
2.0X
3.7X
1.2X
1.0X
1.2X
1.1X
1.2X
2.3X
Ancestral
XBB.1.5
Day 0
Day 28

Fig. S4a

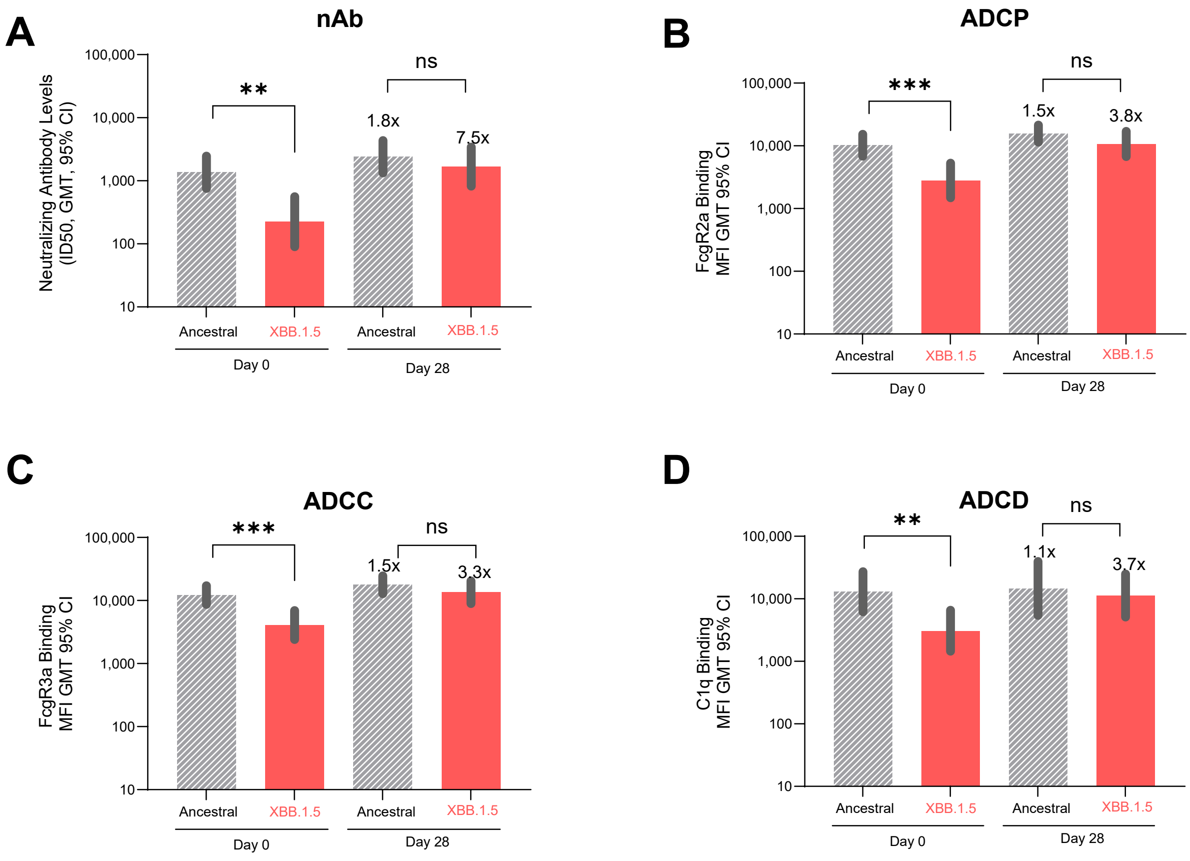
A
nAb
Neutralizing Antibody Levels (ID50, GMT, 95% CI)
100,000
10,000
1,000
100
10
**
ns
1.8x
7.5x
Ancestral
XBB.1.5
Ancestral
XBB.1.5
Day 0
Day 28
B
ADCP
FcgR2a Binding MFI GMT 95% CI
***
ns
1.5x
3.8x
C
ADCC
FcgR3a Binding MFI GMT 95% CI
***
ns
1.5x
3.3x
D
ADCD
C1q Binding MFI GMT 95% CI
**
ns
1.1x
3.7x

Fig.s4b

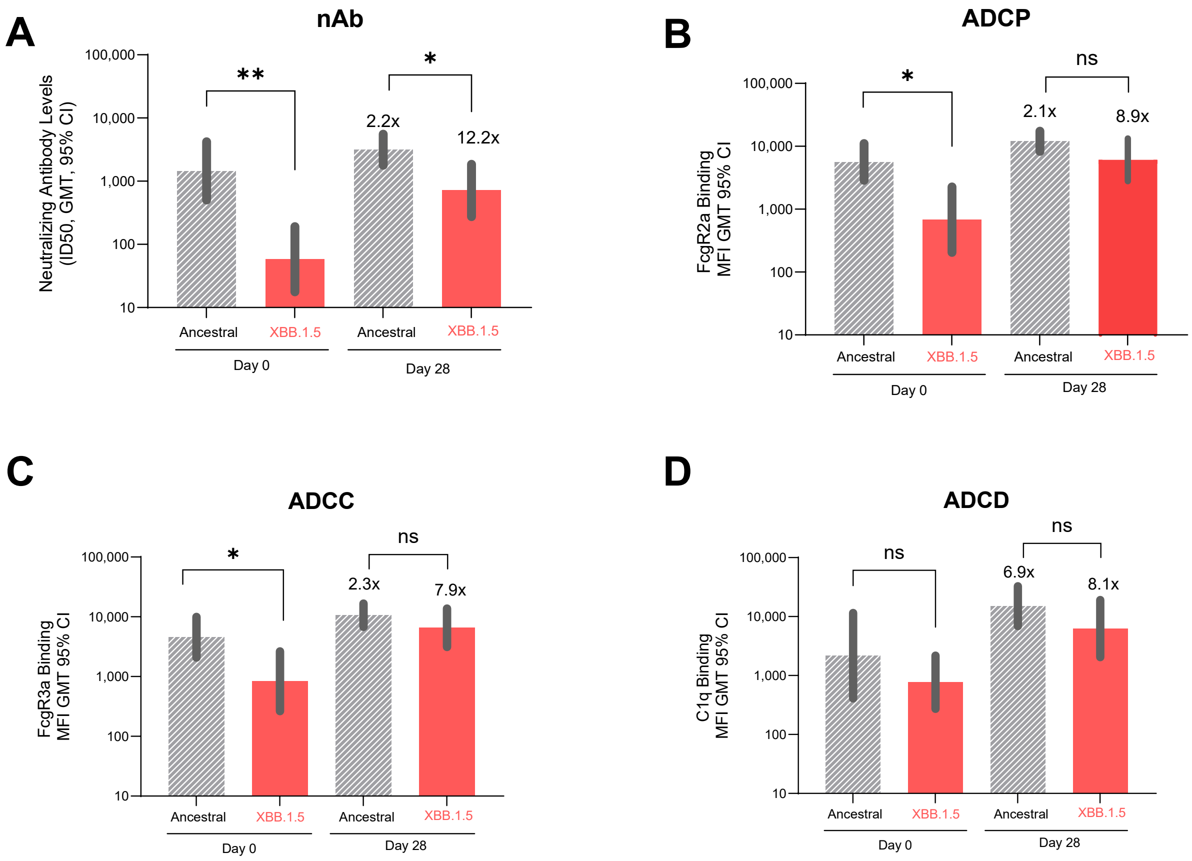

A
nAb
Neutralizing Antibody Levels (ID50, GMT, 95% CI)
100,000
10,000
1,000
100
10
**
*
2.2x
12.2x
Ancestral
XBB.1.5
Ancestral
XBB.1.5
Day 0
Day 28
B
ADCP
FcgR2a Binding MFI GMT 95% CI
100,000
10,000
1,000
100
10
*
ns
2.1x
8.9x
Ancestral
XBB.1.5
Ancestral
XBB.1.5
Day 0
Day 28
C
ADCC
FcgR3a Binding MFI GMT 95% CI
100,000
10,000
1,000
100
10
*
ns
2.3x
7.9x
Ancestral
XBB.1.5
Ancestral
XBB.1.5
Day 0
Day 28
D
ADCD
C1q Binding MFI GMT 95% CI
100,000
10,000
1,000
100
10
ns
ns
6.9x
8.1x
Ancestral
XBB.1.5
Ancestral
XBB.1.5
Day 0
Day 28

Fig.s4c

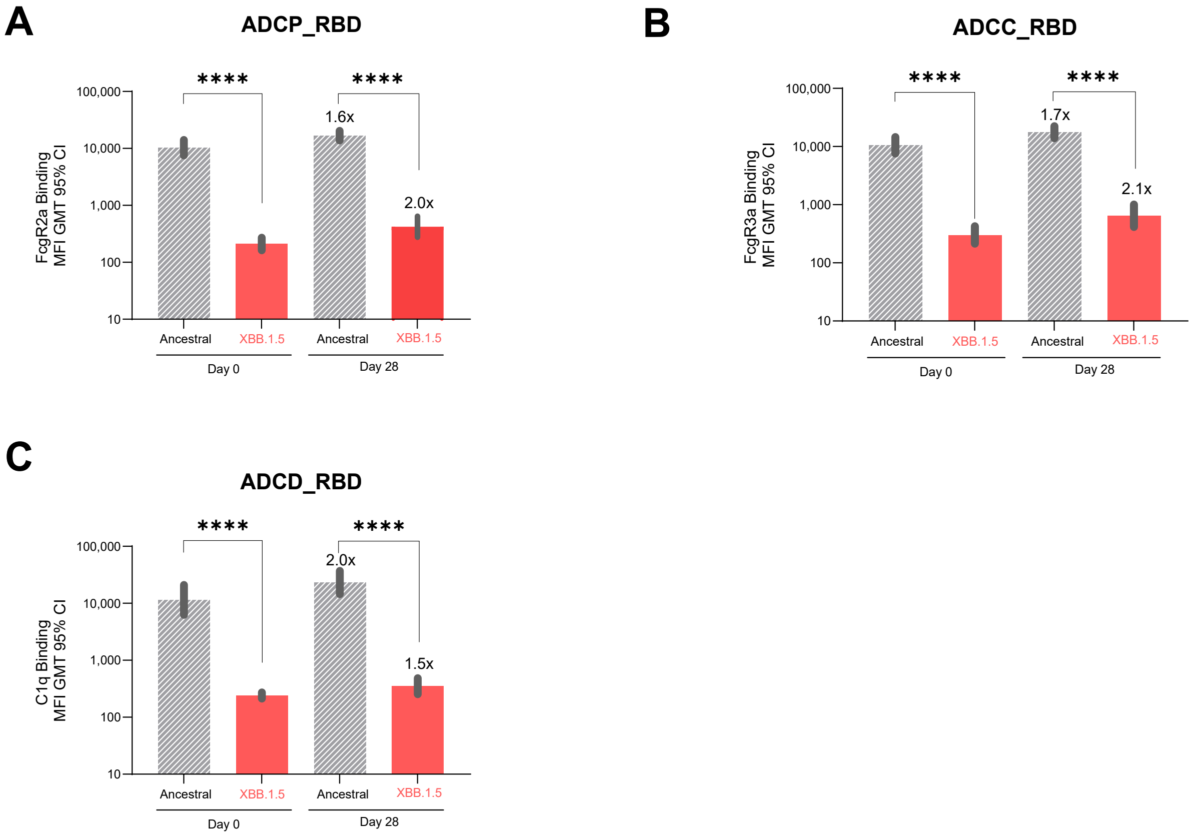
A
ADCP_RBD
FcgR2a Binding
MFI GMT 95% CI
100,000
10,000
1,000
100
10
****
****
1.6x
2.0x
Ancestral
XBB.1.5
Ancestral
XBB.1.5
Day 0
Day 28
B
ADCC_RBD
FcgR3a Binding
MFI GMT 95% CI
100,000
10,000
1,000
100
10
****
****
1.7x
2.1x
Ancestral
XBB.1.5
Ancestral
XBB.1.5
Day 0
Day 28
C
ADCD_RBD
C1q Binding
MFI GMT 95% CI
100,000
10,000
1,000
100
10
****
****
2.0x
1.5x
Ancestral
XBB.1.5
Ancestral
XBB.1.5
Day 0
Day 28

Fig.s4d

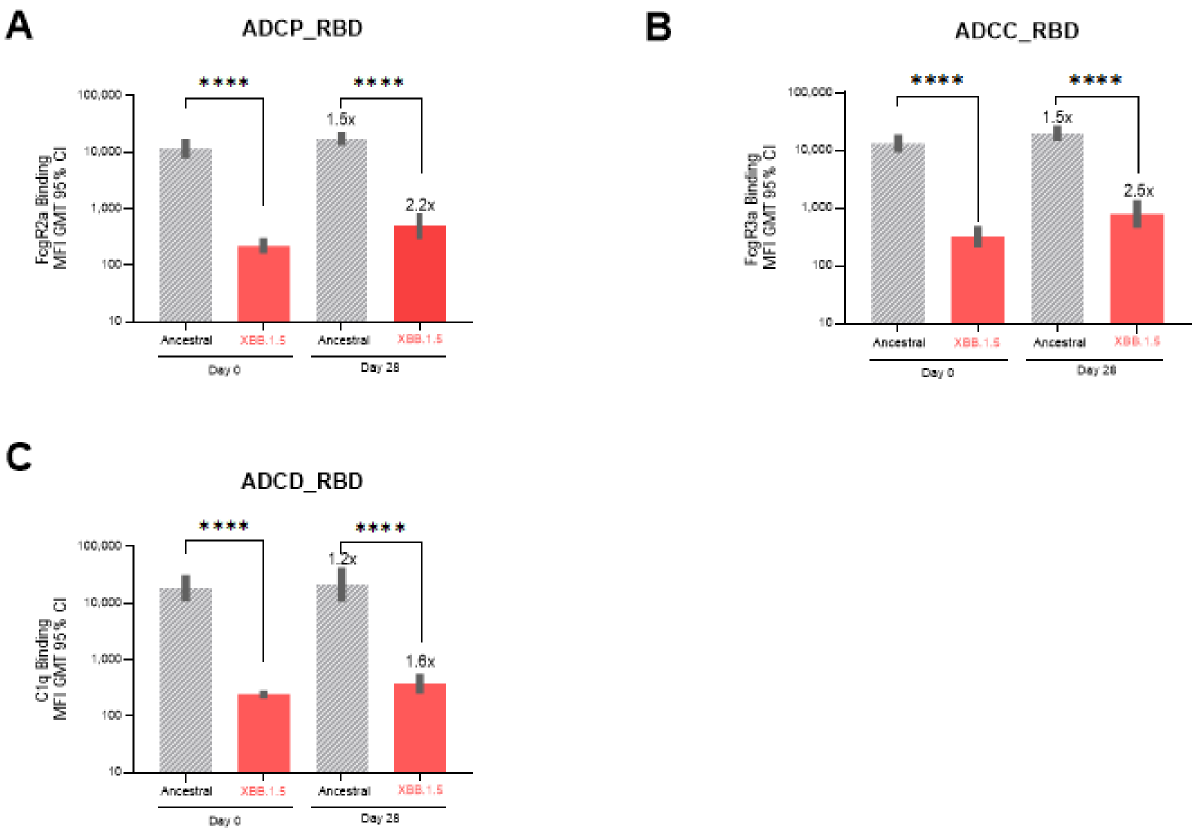

A
ADCP_RBD
FcgR2a Binding
MFI GMT 95% CI
100,000
10,000
1,000
100
10
****
****
1.5x
2.2x
Ancestral
XBB.1.5
Ancestral
XBB.1.5
Day 0
Day 28
B
ADCC_RBD
FcgR3a Binding
MFI GMT 95% CI
****
****
1.5x
2.5x
C
ADCD_RBD
C1q Binding
MFI GMT 95% CI
****
****
1.2x
1.6x

Fig.s4e

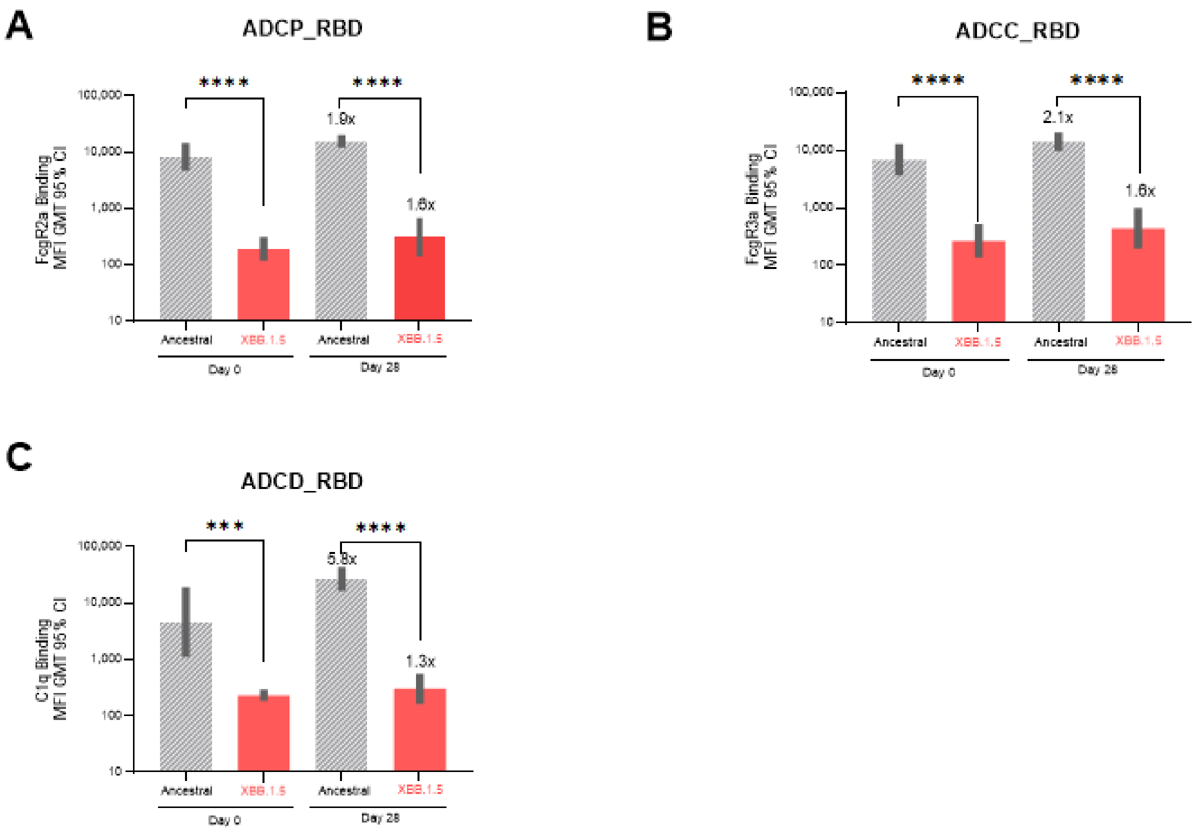

A
ADCP_RBD
FcgR2a Binding
MFI GMT 95% CI
100,000
10,000
1,000
100
10
****
****
1.9x
1.6x
Ancestral
XBB.1.5
Ancestral
XBB.1.5
Day 0
Day 28
B
ADCC_RBD
FcgR3a Binding
MFI GMT 95% CI
100,000
10,000
1,000
100
10
****
****
2.1x
1.6x
Ancestral
XBB.1.5
Ancestral
XBB.1.5
Day 0
Day 28
C
ADCD_RBD
C1q Binding
MFI GMT 95% CI
100,000
10,000
1,000
100
10
***
****
5.8x
1.3x
Ancestral
XBB.1.5
Ancestral
XBB.1.5
Day 0
Day 28
